# Multi-scale integration of human brain vascular and CSF proteomes reveals biomarkers of cerebral amyloid angiopathy linked to Alzheimer’s disease risk

**DOI:** 10.1101/2025.10.08.25337413

**Authors:** Aleksandra M. Wojtas, Saima Rathore, Eric B. Dammer, Lei Yu, Fatemeh Seifar, Joshna Gadhavi, Anantharaman Shantaraman, Duc M. Duong, Fang Wu, Yue Liu, Adam N. Trautwig, Edward J. Fox, Kaylor M. Kelly, Marla Gearing, Srikant Rangaraju, Sharham Oveisgharan, Gareth R. Howell, Julie A. Schneider, Edward B. Lee, Vilas Menon, David A. Bennett, James J. Lah, Todd E. Golde, Erik C.B Johnson, Zachary T. McEachin, the Alzheimer’s Disease Neuroimaging Initiative, Aliza P. Wingo, Thomas S. Wingo, Allan I. Levey, Nicholas T. Seyfried

## Abstract

Cerebrovascular disease frequently co-occurs with amyloid-β (Aβ) plaques and tau tangles, the pathological hallmarks of Alzheimer’s disease (AD), compounding cognitive decline. Aβ deposition is also central to cerebral amyloid angiopathy (CAA), yet peripheral biomarkers that specifically reflect CAA-related vascular pathology remain elusive. Here, we integrated proteomic, imaging, clinical, neuropathological, and genomic data from brain and cerebrospinal fluid (CSF) to define molecular signatures distinguishing CAA from plaque pathology. Proteomic profiling of 118 cerebrovascular-enriched brain samples quantified over 11,000 proteins, revealing co-expression modules enriched for extracellular matrix (ECM) components strongly linked to CAA. In CSF proteomes from 1,104 individuals, vascular ECM module proteins (e.g., CRIP1, LTBP1, PRSS23) were associated with CAA, white matter hyperintensities (WMH), microbleeds, and infarcts but not Aβ plaque burden. Integrating AD genome-wide association studies and CSF protein quantitative trait loci (pQTLs) identified CRIP1 as a candidate causal protein for AD. Carriers of the minor allele at the CRIP1 pQTL exhibited reduced CRIP1 levels, less WMH, and lower CSF levels of ECM proteins linked to CAA. In vitro, CRIP1 bound Aβ and accelerated fibril formation, providing a mechanistic link to vascular amyloid pathology. These findings establish overlapping brain and CSF biomarkers for CAA and identify CRIP1 and vascular ECM pathways as candidate targets for precision diagnostics and therapeutic intervention.

## Introduction

Vascular contributions to cognitive impairment and dementia (VCID) encompasses a broad spectrum of cerebrovascular pathologies, including large vessel atherosclerosis, small vessel disease (SVD), microinfarcts, white matter lesions (WML), and cerebral microbleeds, that collectively drive cognitive decline and pose a critical challenge for aging populations worldwide^1-3^. Cerebrovascular disease commonly co-exists with amyloid-β (Aβ) plaques and tau tangles, pathological hallmarks of Alzheimer’s disease (AD), each synergistically exacerbating cognitive impairment. A central manifestation of cerebrovascular involvement in AD is cerebral amyloid angiopathy (CAA)^4,5^, characterized by Aβ deposition in vessel walls, leading to vessel fragility, microbleeds, and increased risk of hemorrhagic stroke^6^. Moreover, vascular dysfunction in CAA exacerbates white matter disease and impairment of cerebral blood flow^7^, potentially contributing to dementia through mechanisms distinct from those caused by amyloid plaques. Understanding this divergence has become particularly important in the context of anti-Aβ immunotherapies. While these therapies aim to reduce amyloid plaques in the brain, they have been linked to worsening CAA-related complications, such as amyloid-related imaging abnormalities (ARIA), including cerebral edema or hemorrhage in a subset of AD patients undergoing treatment^8–10^. This dual impact highlights the urgency for accurate distinguishment between amyloid pathologies to improve our understanding of vascular contribution to dementia and pave the way for more effective and individualized therapeutic strategies.

A data-driven, proteomic network-based approach has emerged as a powerful strategy in dissecting the complexity of AD pathophysiology in the brain, allowing for the identification of novel biological pathways and disease-relevant modules that are highly reproducible across independent patient cohorts^11,12^. Moreover, integrating AD brain-derived network analyses with proteomic profiling of cerebrospinal fluid (CSF) has provided valuable insights into AD heterogeneity, facilitating the discovery of key biomarkers and potential therapeutic targets^11,13,14^. Recent studies^15,16^, including our own^17^, have characterized the proteomic landscape of human cerebrovasculature in AD, revealing substantial alterations in matrisome proteins associated with amyloid accumulation in brain vasculature. However, since Aβ deposition is a shared pathological hallmark of both AD and CAA^4,5^, identifying peripheral biomarkers that specifically reflect CAA-related vascular abnormalities remains a significant challenge. Magnetic resonance imaging (MRI) has revolutionized the diagnostic approach to CAA, with the Boston Criteria providing a framework that leverages MRI-visible features such as presence of lobar cerebral microbleeds, white matter hyperintensities (WMH), infarcts, and intracerebral hemorrhages in living patients^18–20^. Thus, an alignment of MRI findings with a pattern of amyloid distribution in the cerebrovasculature represents a unique opportunity for more comprehensive and mechanistic understanding of CAA and its differentiation from amyloid plaque pathology.

In the current study we aimed to define protein biomarkers that are selectively associated with CAA, independent of amyloid plaques, by integrating vascular-enriched proteomics of postmortem AD brain tissue with CSF proteomics, and neuroimaging. Generating the most comprehensive cerebrovascular proteome to date, with over 11,000 quantified proteins, we identified modules of co-expressed proteins across 118 AD and control samples that significantly map to diverse biological pathways implicated in vascular dysfunction and CAA. An independent, proteome-wide correlation analysis of brain tissues from 887 cases in the Rush Religious Orders Study and Memory and Aging Project (ROSMAP), each with well-defined pathological measures, unbiasedly identified four cerebrovascular modules enriched for CAA markers. Alterations in the extracellular matrix (ECM) emerged as a central theme, enabling the prioritization of proteins most strongly associated with CAA. The specificity of these modules to CAA, distinct from parenchymal amyloid, was further supported by localized proteomic approach of micro-dissected vascular Aβ deposits. CSF proteomics and high-resolution MRI data from 1,104 participants of the Alzheimer’s Disease Neuroimaging Initiative (ADNI) revealed that ECM proteins such as LTBP1, CRIP1, C1QC, PRSS23, ELN, and CEMIP were associated with vascular imaging abnormalities and neuropathological features of CAA, but not with PET AV45 signal, highlighting their independence from Aβ plaques. Proteogenomic analysis of ADNI CSF samples further identified a genetic variant in *CRIP1* (rs35590487) associated with its protein level and WMH in a dose-dependent manner independent from apolipoprotein E4 (*APOE4)* risk. We further showed that CRIP1 binds Aβ and accelerates Aβ fibril formation providing a potential mechanistic link to the vascular amyloid pathology in CAA. Together, these findings define molecular signatures of CAA across brain and CSF, offering a foundation for precision diagnostics and therapeutics targeting vascular contributions to cognitive impairment in AD.

## Results

### Extracellular matrix proteins are altered in AD cerebrovasculature

Characterizing the proteome of isolated cerebrovasculature is crucial for uncovering the unique molecular architecture of vascular cells, which are often obscured in bulk brain proteomics due to the predominance of neuronal and glial-specific proteins. This approach is particularly critical for studying CAA, as it enables precise identification of vascular-specific protein alterations driven by amyloid-associated pathways. To investigate molecular mechanisms underlying vascular pathology in AD, we performed tandem mass tag (TMT)–based quantitative proteomics on 118 individually isolated cerebrovascular fractions from postmortem human brain tissue from the Emory Brain Bank (Emory) and the University of Pennsylvania School of Medicine Brain Bank (UPenn) (**Figure 1**). The neuropathological assessment of amyloid in brain parenchyma (CERAD score) and vasculature (CAA) as well as tau accumulation (Braak staging) in the brain guided the classification of cases as neurologically healthy controls (Emory: N=20; UPenn: N=25) or Alzheimer’s disease (Emory: 61; UPenn: N=12). Frontal cortex was selected given its vulnerability for pathological accumulation of amyloid in brain parenchyma and blood vessels^21^. Samples were matched for age, sex, and neuropathological traits (**Table S1**). As previously described^17^, we extracted brain vasculature using dextran density centrifugation approach followed by the enzymatic digestion, TMT labeling, and liquid chromatography tandem MS (TMT-MS) (**Figure 1**).

**Figure 1.**
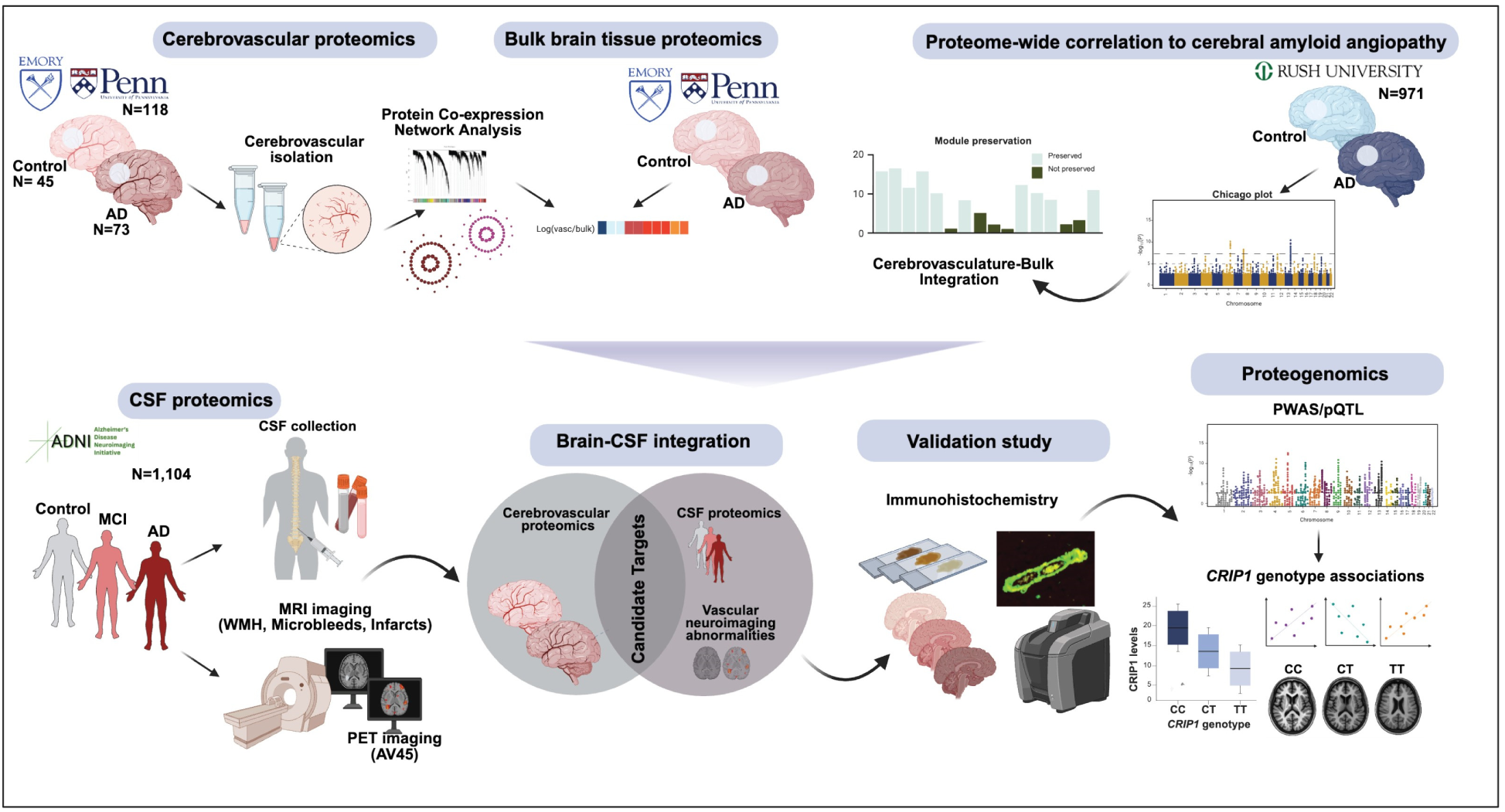
Multi-modal proteomic and imaging integration identifies proteins associated with CAA pathology. Schematic overview of the study design. Human cerebrovascular fractions were isolated from postmortem cortical tissue of AD and control cases, followed by enzymatic digestion and tandem mass tag mass spectrometry (TMT-MS). Corresponding bulk brain proteomes were used to identify modules enriched in vascular-specific proteins. Integration of the cerebrovascular protein network with large-scale bulk brain proteomic data from 887 individuals in the ROSMAP cohort enabled prioritization of brain modules specifically associated with CAA pathology. CSF proteomic profiles from 1,104 ADNI participants, spanning cognitively normal controls, mild cognitive impairment (MCI), and AD, were linked to vascular neuroimaging features and mapped back to the cerebrovascular proteome. Candidate proteins were validated by immunohistochemistry in brain tissue, revealing markers associated with CAA and vascular lesions. Proteogenomic analysis of 988 ADNI CSF samples, incorporating genome-wide association study (GWAS) data and protein quantitative trait loci (pQTL) mapping, identified CRIP1 as a candidate causal protein. Both CSF CRIP1 levels and its cis-pQTL (rs35590487) were significantly associated with white matter hyperintensity (WMH) burden.

To globally characterize proteomic signature of vascular enriched fractions, we assessed contribution of main brain cell types to vascular proteomes using previously published cell-specific marker list^22^. As expected, we observed a pronounced enrichment of endothelial-specific proteins compared to neurons and glial cells (**Figure S1A**). Given some degree of heterogeneity in sample fractionation, we normalized the proteomic data for endothelial specific signatures across control and AD brain tissues (**Figure S1**). Using a well-established data processing pipeline, we further employed batch correction analysis (**Figure S1B**) and adjustment for confounding effects of age and postmortem interval (PMI) (**Figures S1C** and **S1D**).

The resulting cerebrovascular proteome, comprising of 11,408 total quantified proteins, was used to generate a protein co-expression network using the weighted correlation network analysis (WGCNA) algorithm (**Figure 2A**). The constructed cerebrovascular network organized proteins into 76 modules with biologically meaningful expression patterns across control and diseased samples and ranked them according to the module size (**Table S2**). Top ranking Gene Ontology (GO) terms, encompassing biological function, process, and components, were considered in determining principal biology of each network module (**Table S3**). Module composition revealed associations with various brain cell types (**Table S4**), particularly vascular cells linked to ECM organization and remodeling, circulation, and immune response (**Figure 2A**). Moreover, 60% of modules showed positive or negative correlation with disease and substantial correlation with AD neuropathological hallmarks, including amyloid deposition in the cerebrovasculature (e.g. CAA) or plaque in the brain parenchyma (CERAD), and tau staging (Braak) (**Figure 2A**). Interestingly, we found 14 modules significantly linked to race and 19 modules with a significant association to sex (*p value<0.05*) (**Figure 2A**). Comparative analysis of module eigenproteins, which represents the first principal component of the module expression level, across African American and non-Hispanic White individuals revealed significantly elevated expression (*p value<0.05*) of extracellular matrix (ECM) (M10), actin cytoskeleton (M25), and synaptic function (M63, M28) modules, alongside decreased RNA processing and translation modules (M12, M53, M18) (**Figure S2A**). Differential abundance analysis revealed a strong enrichment of extracellular matrix proteins (*p value <0.05*), particularly those mapping to module M37, across diverse populations (**Figures S2B-S2D**, **Table S5** and **Table S6**). In addition, the levels of proteins associated with AD, including MAPT and Aβ fragments (Aβ42 and Aβ40), were largely consistent across racial groups, aligning with our findings from the bulk brain proteome^23^ (**Figure S2E**). These observations mirror previous studies that showed no difference in CAA prevalence in African Americans with AD compared to non-Hispanic White individuals^24,25^ and therefore unlikely to account for racial disparities in AD susceptibility. Additionally, among modules with a significant alteration in AD, males showed higher levels of module eigenprotein represented by collagen organization (M7) and synaptic function (M63, M28) compared to females with concomitant decrease in modules linked to immune response and circulatory system development (**Figure S3, Table S7**). In summary, our cerebrovascular proteomic network outlines interrelationships between disease status, neuropathological traits and demographic features.

**Figure 2.**
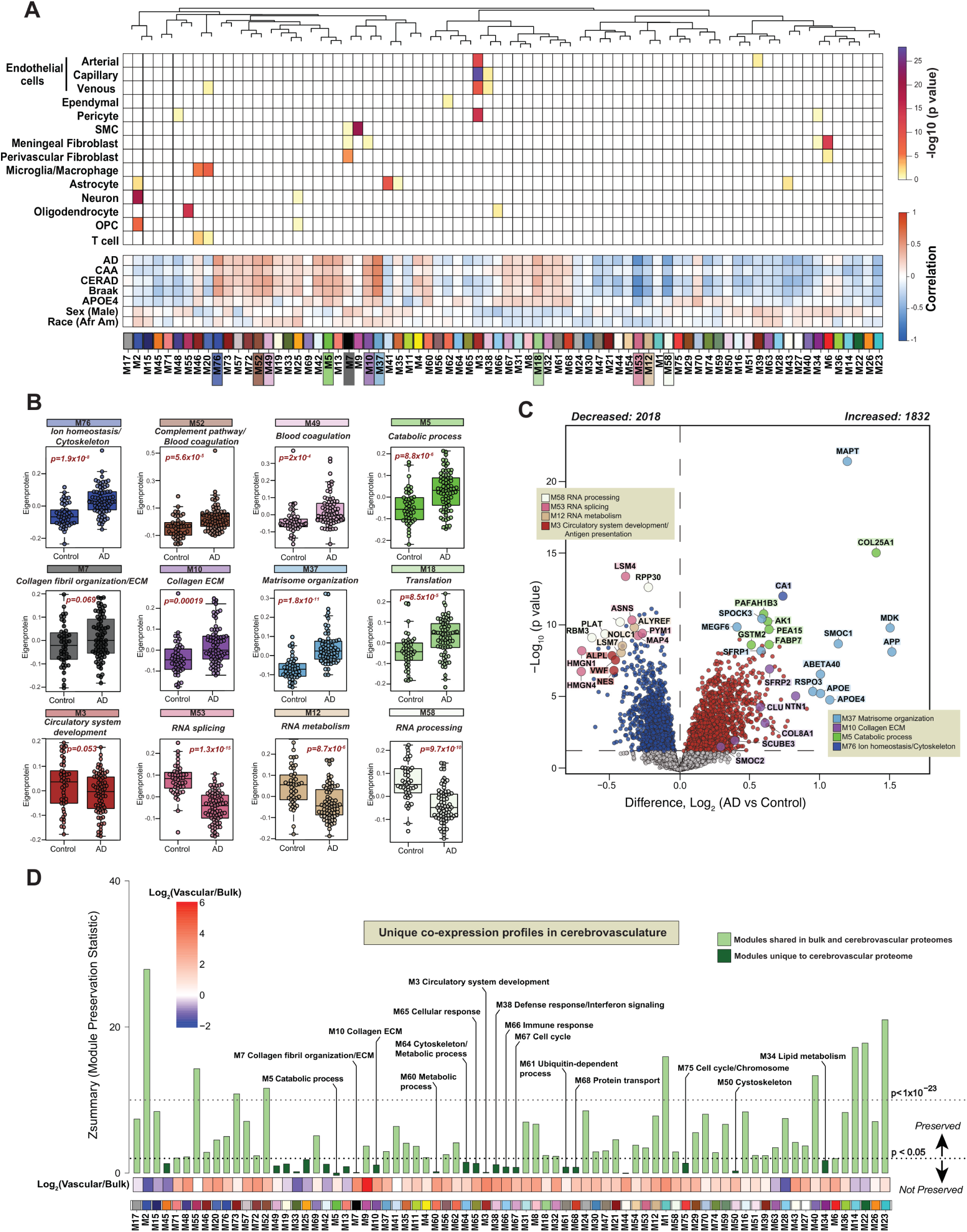
Network-based analysis of cerebrovascular proteome identifies modules associated with cell types, AD pathology and CAA. (A), A protein co-expression network was constructed from 11,408 proteins using the WGCNA approach and consisted of 76 protein co-expression modules (each module represented by a different color and number for decreasing rank size). Modules are organized based on their relatedness and shown with a dendrogram. The association of brain cell types with protein modules was assessed in 14 cell types by hypergeometric overlap of each module’s gene products with the cell type specific marker list for each brain cell type de novo extracted from single-nuclei RNA-seq data and one-sided Fisher’s exact test with Benjamini-Hochberg correction was employed. Scale bar indicates cell type enrichment (from yellow modestly significant to darker brown/purple indicating stronger enrichment). Module eigenproteins were correlated with binary disease/control status, AD-associated pathological hallmarks (CAA, CERAD, Braak), *APOE4* dose, sex, and race. Two-color heatmap represents strength of positive (red) or negative (blue) correlation, with *P* values provided for all correlations with *P* < 0.05. (B), Module eigenprotein (ME) levels by diagnosis status (Controls, N = 45; AD, N=73) were presented for selected protein modules of interest. P values are provided for each module and were obtained by Student’s t-test. Box plots represent median, 25^th^ and 75^th^ percentile while box whiskers encompass actual data points up to 1.5 times the nearest interquartile range. Representative biological process of each module was assessed by gene ontology (GO) analysis. (C), Volcano plot illustrating differential abundance of proteins between control and AD subjects. The x axis represents the log_2_ fold change (AD vs Control) while the y axis shows the -log_10_ statistical *p* value calculated for all proteins in each group. *P* values were obtained from Student’s t-test Proteins significantly increased in AD vs control are shown in red, whereas proteins significantly decreased in AD are highlighted in blue. Proteins with unchanged levels are represented in grey. Proteins of interest are shown as enlarged dots and shaded according to color of the module membership. (D) Module preservation of the TMT cerebrovascular network into the ROSMAP TMT bulk protein network. Modules that had a Z_summary_ score greater than or equal to 1.96 (or p< 0.05) were considered to be preserved, whereas modules that had a Z_summary_ score greater than or equal to 10 (or p< 1×10^−23^) were considered to be highly preserved. Modules that had a preservation Z_summary_ score less than 1.96 were not considered to be preserved. Each bar represents a protein module and is shaded according to color of its preservation status. Mean log_2_ ratio of vascular to bulk proteomic abundance was performed to identify modules enriched in vascular proteome, red indicates enrichment and blue indicates depletion.

To deepen our understanding of interplay between AD vascular modules and neuropathological changes, we assessed the global levels of module eigenprotein across control and disease brains (**Figure 2B**). Specifically, a cluster of 13 modules, associated with ECM function (M10, M37), coagulation/immune biology (M73, M57, M72, M52, M49), metabolism (M76, M5, M19, M13), actin cytoskeleton (M25) and protein folding (M42) were significantly elevated in AD (*p value<0.05*) and positively correlated with pathological accumulation of amyloid and tau in the brain (**Figures 2A** and **2B**). Conversely, a set of 16 modules exhibited a significant reduction (*p value<0.05*) in AD and was primarily associated with transport (M42, M51), mitochondrial function (M16, M23), and RNA processing and translation (M21, M44, M53, M12, M58, M29, M40), among others with dysregulation of transcription and RNA metabolism previously reported in recent localized proteomics study^16^ (**Figures 2A** and **2B**). Pairwise differential abundance analysis revealed widespread proteomic shifts in the cerebrovascular proteome in AD, with 1,832 (1144 with ≤ 5% FDR) proteins significantly increased and 2,018 (1214 with ≤5% FDR) decreased compared to controls (*p value<0.05*, **Figure 2C, Table S8**).

Amyloid-associated proteins with high specificity in discriminating AD from other proteinopathies^12,17^ (SMOC1, MDK, APOE, SFRP1, SMOC2, NTN1) were enriched in ECM-driven modules M10 and M37. M10 (Collagen ECM) encompassed 245 proteins involved in matrix structure, angiogenesis, vascular permeability, and Wnt signaling highlighting its role in vascular remodeling. M37 (Matrisome organization), composed of 98 co-expressed proteins, also showed robust enrichment of amyloid- and tau-related proteins (APP, MAPT and Aβ40 and Aβ42 amyloid fragments), underscoring its relevance to AD pathology (**Figure 2C**). In addition, proteins involved in coagulation and immune responses were markedly elevated in AD vasculature compared with controls. In contrast, proteins reduced in AD were predominantly involved in gene regulation, RNA processing, splicing, and mitochondria function reflecting broader disruptions in neuronal homeostasis (**Figure 2C**). Together, these findings define a distinct AD cerebrovascular proteomic signature, characterized by ECM remodeling and immune process, mirroring and expanding upon changes seen in brain parenchyma.

### Extracellular matrix modules associated with AD pathologies are not well preserved in the bulk proteome

Drawing from our earlier discoveries that bulk proteomic analyses fail to capture the complexity of AD-associated vascular changes^17^, we employed network preservation statistics to assess how cerebrovascular protein modules are represented in bulk brain tissue (**Figure 1**). Using co-expression networks from 971 dorsolateral prefrontal cortex (DLPFC) samples from the ROSMAP cohort (**Table S9**), we found that 54 vascular modules were preserved in the bulk proteome, with 9 showing strong preservation (Zsummary > 10, q < 1×10⁻²³), including modules related to transcription, synaptic function, mitochondrial activity, and immune signaling (**Figure 2D**). An additional 45 modules showed moderate preservation (Zsummary > 1.96, q < 0.05), spanning processes such as ion homeostasis, lipid metabolism, and ECM organization (**Figure 2D**). Importantly, 23 modules exhibited cerebrovascular-specific co-expression pattern not preserved in the bulk network. These unique modules encompassed biology associated with vascular-specific functions (e.g., M3, M7, M10, and M49), immune activation (M38, M44, M65, and M66), metabolism (M13, M19, M33, M34, M45, and M60), and cell cycle regulation (M67 and M75), with nearly half showing significant alterations in AD, strong associations with neuropathological burden, and enrichment in cerebrovasculature (**Figure 2D**). Interestingly, the divergence of ECM modules (M10 and M37) in the current network was underscored by their distinct preservation pattern. Although M10 was enriched with known amyloid-associated proteins (e.g. CLU, NTN1, SPON1) the co-expression signature reflected vascular-specific function. In contrast, M37, highly homologous to M42 (Matrisome) in our previous bulk proteomic studies^12^, showed shared co-expression profile in bulk and isolated vasculature, potentially indicating broader involvement across vascular and parenchymal compartments. We further validated cerebrovascular network robustness by comparing to an independent cerebrovascular proteome from AD, PSP, and control frontal cortex samples (N=62 isolated cerebrovascular fractions)^17^, revealing ∼97% module preservation and confirming the reproducibility of vascular-specific networks across disease contexts (**Figure S4**).

### APOE shows strong association with ECM modules

The allelic variation in apolipoprotein E (*APOE*) has shown the strongest susceptibility for sporadic AD and CAA, with the presence of the *APOE ε4* allele increasing risk in a dose dependent manner^26,27^. APOE4 accelerates breakdown of the BBB^28,29^ and increases risk for cognitive impairment during normal aging^29^. Furthermore, it has been reported that APOE4 is associated with disruption of the perivascular drainage pathway, an effect mediated by APOE-related alterations of basement membrane proteins^30,31^. However, the underlying molecular mechanisms of this genetic susceptibility on cerebrovascular function are still poorly understood. The APOE4 isoform, marked by a cysteine-to-arginine substitution, can be uniquely detected via mass spectrometry using a tryptic peptide (LGADMEDVR). We previously quantified this peptide across over 1,100 brain tissue samples and demonstrated >8-fold enrichment in APOE *ε4* carriers compared to non-carriers, with negligible signal in non-carriers likely due to genotyping errors or technical artifacts^23^. In this study, we performed a proteome-wide correlation analysis of bulk DLPFC tissue from 873 ROSMAP cases to identify proteins associated with *APOE4* genotype. We identified 27 proteins positively associated with APOE4, with the most significant changes found in ECM-associated modules M10 (e.g., NTN1, HTRA1, CLU) and matrisome module M37 (e.g., APOE4, MDK, SMOC1, APP, SLIT2, SPOCK3) (**Figure S5A, Tables S9** and **S10**), in line with previous reports^32^. APOE4 protein itself emerged as the most strongly associated protein, validating our approach. Conversely, eight proteins showed a negative association with APOE4, many of which are involved in axonal biology and intracellular transport (e.g., RTN4, STAM, KIF5C, DOCK3, PTPN11) (**Figure S5A**). Two modules (M10 and M37) in our cerebrovascular network were identified as significantly enriched with proteins conferring association with APOE4 (**Figure S5B, Table S11**). Additionally, the eigenprotein levels of M10, M37, and M5 exhibited a strong bias towards the increase with an increasing dose of *ε4* allele (**Figure S5C**) highlighting their biological relevance to APOE4-driven pathology.

### Divergence of ECM modules in response to vascular and parenchymal amyloid pathology

To unbiasedly nominate proteins associated with CAA, we conducted a series of proteome-wide correlation analyses using previously established methods on large bulk proteomic datasets^32^ (**Figure 1**). Logistic regression models were performed on the 10,030 proteins in the ROSMAP proteome across 887 individual cases (**Table S9**) using a semiquantitative measure of CAA burden (0-3 in severity scale), as the outcome. Twenty proteins survived Bonferroni correction (i.e., p < 0.05/10000), with positive -log10 p values indicating a positive association with CAA following adjustments for neuritic plaque burden (**Figure 3A**) or tau tangles (**Figure 3B**). The overwhelming majority of CAA-associated proteins were strongly representative of ECM biology and predominantly associated with M10 and M37 (**Figure 3, Table S12**).

**Figure 3.**
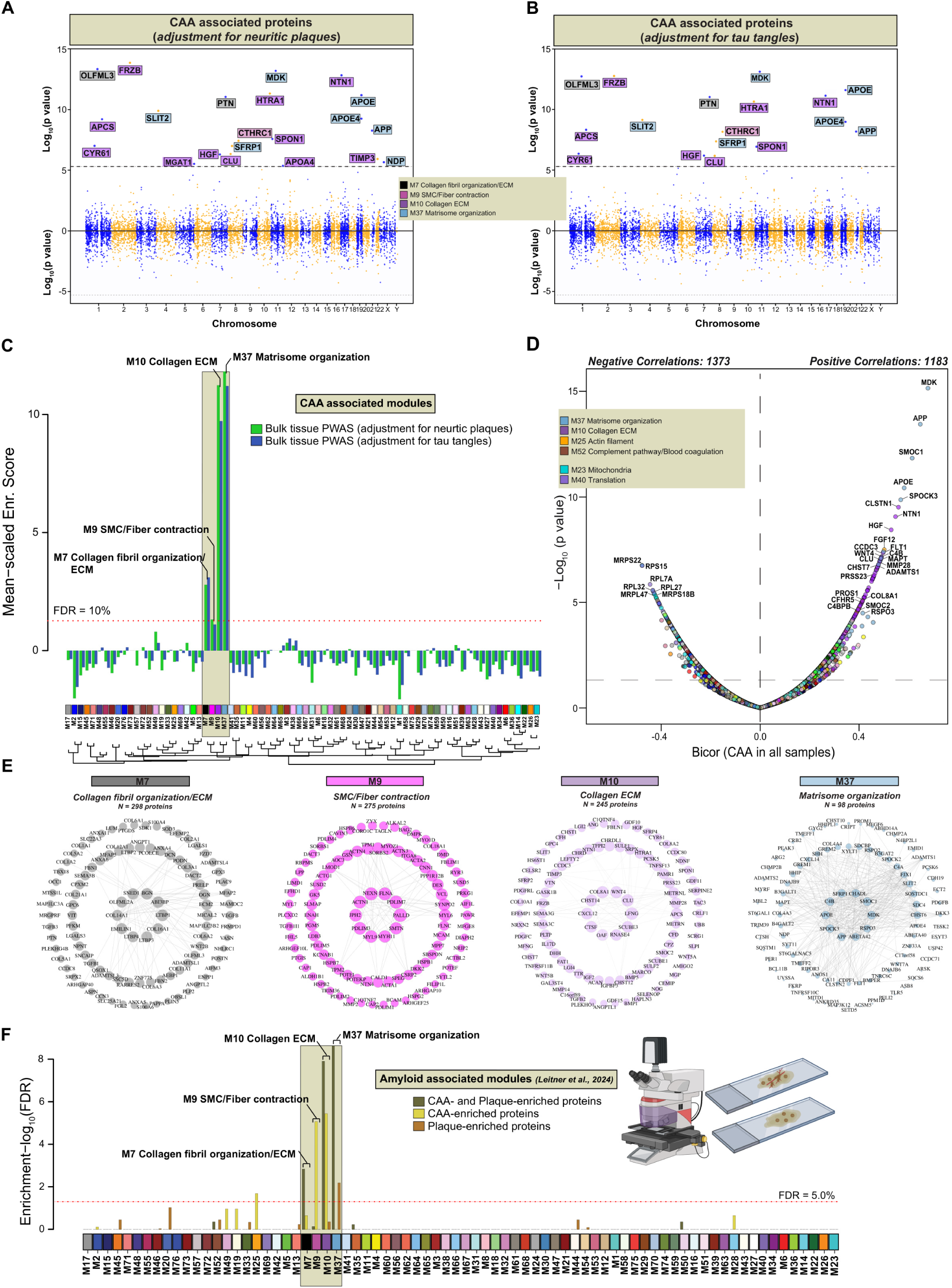
Proteome-wide correlation analyses and vascular network integration reveal extracellular matrix (ECM) modules enriched for proteins linked to CAA. (A-B) Individual measured proteins (n=10,030) organized by chromosome location (x-axis) highlighted in the Chicago plots survived Bonferroni correction (i.e., p < 0.05/10000), with positive -log10 p values (y-axis) indicating a positive association with CAA following adjustments for neuritic plaque (A) or tau tangle (B) burden. Logistic regression was performed in the ROSMAP bulk proteome across 887 individual cases using a semiquantitative measure of CAA burden (0-3 in severity scale) as the outcome. The dotted lines above and below line of zero indicate the levels above and below which the associations are statistically significant after Bonferroni correction. The significant associations above the zero line indicate upregulated proteins in relation to CAA. (C) CAA associated proteins (A-B) were mapped to cerebrovascular modules. The horizontal dotted lines indicate the thresholds for permutation test false discovery rate (FDRs of 10%) above which the enrichment of CAA associated proteins was considered significant. The yellow box highlights four ECM modules with significant enrichment of CAA associated proteins (M7, M9, M10, M37). (D) Biweight midcorrelation (Bicor) volcano plot illustrating correlation of cerebrovascular proteins to CAA. The x-axis represents the bicor correlation with CAA in AD and controls while the y-axis shows the -log_10_ statistical *p value* calculated for all proteins. 1183 proteins showed a significant positive correlation to CAA, whereas 1373 proteins showed a significant negative correlation to CAA. Proteins are colored based on their module membership. (E) iGraphs showing top ∼100 proteins identified in M7, M9, M10, and M37 modules. The colors represent the module membership. Lines between proteins represent topological overlap matrix weight corresponding to the similarity of correlated patterns of node pairs in the cerebrovascular network. (F) Integrated approach of localized proteomic dataset with cerebrovascular network. The horizontal red line indicates the threshold for permutation test false discovery rate (FDRs of 5%) above which the enrichment of proteins correlated with amyloid-related proteins was considered significant. Yellow box outlines modules: M7, M9, M10, and M37 with significant association to CAA.

To expand upon these findings, we integrated the results from bulk brain proteome-wide correlation analysis of CAA adjusted for AD core pathologies, with our independent cerebrovascular proteomic network. Through this approach, we identified four modules that showed significant enrichment of proteins conferring a positive association with CAA: M7 (Collagen fibril organization/ ECM), M9 (SMC/Fiber contraction), M10 (Collagen ECM), and M37 (Matrisome organization) strengthening validity of our observations (**Figure 3C** and **Table S13**).

We next distinguished proteins with significant positive (N=362) and negative (N=497) correlations to CAA in cerebrovascular proteome (*p value<0.05*) (**Table S14**). Consistently, we observed that protein constituents of ECM modules: M10 (CLSTN1, NTN1, HGF, WNT4, CLU, PRSS23) and M37 (MDK, APP, SMOC1, APOE, SPOCK3, FLT1) displayed a significant bias towards the positive correlation with CAA (**Figure 3D**). In contrast, proteins that exhibited negative correlation with CAA across AD and control samples were associated with mitochondrial function (MRPS22, RPL7A, RPL32, RPS15) (**Figure 3D**). While large scale bulk proteomic analysis facilitated identification of modules significantly associated with CAA, it lacked the resolution to differentiate modules linked to vascular and parenchymal amyloid plaque pathology. To address this, we leveraged high-resolution localized proteomic data from laser captured micro-dissected CAA-positive vessels and previously characterized amyloid plaque proteomes enabling the identification of proteins either uniquely enriched in one compartment or shared between them (**Table S15**)^16^. Mapping these proteins onto the cerebrovascular network uncovered divergent associations of ECM modules. Modules M7, M10, and M37 contained proteins present in both CAA and plaques, reflecting overlapping pathways. M9 was enriched almost entirely with proteins unique to CAA, marking a molecular fingerprint of vascular amyloid deposition. Finally, M37 was uniquely concentrated proteins specific to amyloid plaques (**Figures 3E** and **3F, Table S16**). These findings highlight that while ECM remodeling is a feature of both vascular and parenchymal amyloid pathology, distinct ECM modules are enriched in a compartment-specific manner, suggesting that local microenvironmental signals differentially regulate ECM reorganization in CAA versus plaque deposition.

### CSF proteomic signatures of neuroimaging-related cerebrovascular and amyloid pathology reveal distinct biomarker profiles

Studies focusing on the biomarker identification in CSF have profoundly contributed to the AD research by shedding light on disease mechanism and advancing early disease detection and diagnosis^11,13,14,33^. Building on findings from brain proteomics that revealed distinct protein modules associated with either CAA or parenchymal amyloid plaques, we investigated whether the CSF proteome could identify biomarkers that distinguish between these two forms of amyloid pathology. To this end, we examined 2,492 CSF proteins in a cohort of 1,104 participants from the ADNI study, encompassing individuals with MCI, AD as well as cognitively normal controls (**Figures 1**, **4A, S6, S7, Tables S17** and **S18**). A substantial number of participants had baseline MRI scans assessing WMH, microbleeds, and infarcts, allowing us to directly link CSF protein alterations with neuroimaging markers associated with small vessel disease. Differential abundance analysis revealed widespread proteomic alterations associated with cerebrovascular pathology: over 1,100 proteins were linked to WMH, 340 to microbleeds, and more than 460 to infarcts. Across all three vascular lesions, we observed a consistent increase in proteins involved in ECM, vascular structural integrity, angiogenesis, and cell adhesion, and biological processes central to vascular remodeling and amyloid-associated vessel pathology (*p value<0.05*, linear model; **Figures 4B**-**4D, Table S19**). Importantly, each vascular abnormality also displayed a distinct proteomic fingerprint. WMH was uniquely associated with alterations in proteins regulating complement pathway, cytoskeletal remodeling, oxidative stress response, and axonal maintenance, while showing a marked decrease in synaptic, cell adhesion, and neurodevelopmental markers (**Figures 4B** and **S8**). Microbleeds exhibited a proteomic shift toward angiogenesis, cell proliferation and communication, and ECM dynamics, coupled with reductions in lipid metabolism and lysosomal organization (**Figures 4C** and **S8**). Infarcts demonstrated strong increase in protein signaling pathways associated with phagocytosis, cellular senescence, and fiber contraction, alongside decreased expression of proteins related to neuron development and synaptic organization (**Figures 4D** and **S8**). To contextualize the CSF protein signatures with amyloid pathology in the same individuals, we next analyzed [^18^F] florbetapir or PET AV45 imaging data which detects neuritic plaque burden with high specificity and affinity^34,35^ (**Figures 4E, Table S20**). We observed highly elevated levels of SMOC1 (*p value(FDR) = 2.05×10^-^*^52^), APOE4 (*p value(FDR) = 1.49×10^-^*^28^), YWHAZ (*p value(FDR) = 2.06×10^-^*^43^), YWHAE (*p value(FDR) = 5.48×10^-41^*), YWHAG (*p value(FDR) = 3.92×10^-33^*), PEA15 (*p value(FDR) = 4.47×10^-37^*), among others broadly associated with amyloid metabolism, glycolysis, neuronal injury, and synaptic dysfunction (**Figures 4E** and **S8**).

**Figure 4.**
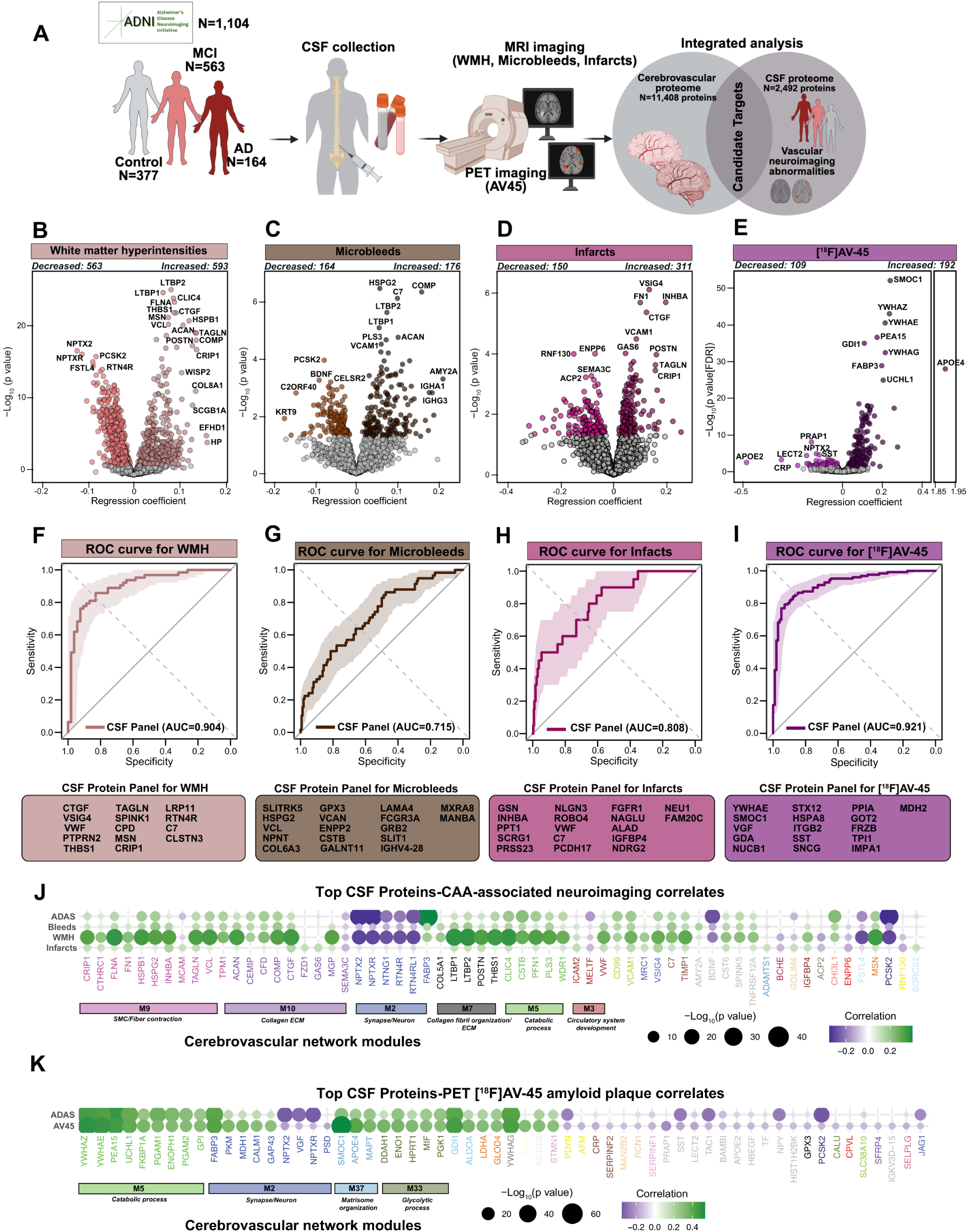
CSF proteomic associations with neuroimaging markers of cerebrovascular pathology and amyloid burden in ADNI. (A) CSF samples were collected from individuals with clinically diagnosed MCI (N=563), AD (N=164) and controls (N=377) from the ADNI cohort. The magnetic resonance imaging (MRI) was conducted on the same set of participants near the baseline. CSF proteins correlated with imaging vascular abnormalities were integrated with cerebrovascular network. (B-E) Volcano plots displaying differential abundance of CSF proteins at P<0.05 correlated with WMH (Increased: 593, Decreased: 563) (B), microbleeds (Increased: 176, Decreased: 164) (C), infarcts (Increased: 311, Decreased: 150) (D) and AV-45 (E) in ADNI participants (WMH: N=775; Infarcts: N=1050; microbleeds: N=732; AV45: N=745; ADAS11: N=1,101). The x axis shows the regression coefficient, the y axis indicates −log 10 statistical p value for all proteins. Select proteins are labeled. (F-I) Support vector machine-based forward feature selection approach was used to model the detection of WMH (top- and bottom tertiles; n=384) (F), microbleeds (G), and infarcts (H). Key proteins associated with the vascular abnormalities are shown in the boxes. The final model trained on the selected proteins of 75% of the data achieved an area under the receiver operating characteristic curve (AUC) of 0.715, 0.808, and 0.904 in predicting microbleeds, infarcts, and white matter hyperintensity, respectively, when evaluated on the left-out 25% of the data. (J) Dot plot highlighting top 62 CSF proteins, selected as the union of the top 30 proteins associated with each of the three imaging-based vascular abnormalities (WMH, microbleeds, infarcts). Each protein is represented by a dot and the protein name is colored by a module membership. The color of each dot indicates correlation with green color showing positive correlation and purple color showing negative correlation. The size of each dot represents −log 10 statistical p value. (K) Dot plot exhibiting top 30 CSF proteins with a positive correlation and top 30 CSF proteins showing a negative correlation with PET AV45. ADAS-11 associations are displayed only for the selected set of proteins both in J and K.

We next assessed the collective power of the CSF proteome in detecting neurovascular imaging manifestations using machine learning. For infarcts and microbleeds, we modeled their presence as a binary variable, with at least one occurrence indicating presence. Our dataset included 194 individuals with microbleeds and 529 without, as well as 81 individuals with infarcts and 969 without (**Figures 4F-4I**). For WMH, we stratified the data based on WMH burden and categorized the top (n=192) and bottom (n=192) tertiles as two distinct classes. Using a support vector machine (SVM)-based forward feature selection approach, we identified key proteins associated with microbleed presence. The most important predictors of microbleeds included SLITRK5 (synapse/axon), HSPG2 and VCL (actin cytoskeleton), NPNT, COL6A3, and GPX3 (collagen fibril organization), VCAN and ENPP2 (myelination), as well as CSTB, GALNT11, LAMA4, FCGR3A, GRB2, SLIT1, IGHV4-28, MXRA8, and MANBA. Similarly, the model identified key proteins associated with WMH and infarcts. For WMH, the most important predictors included VSIG4 (microglia), PTPRN2 and PTN4R (synapse/axon), CRIP1 and TAGLN (SMC/fiber contraction), CTGF (extracellular matrix), as well as VWF, THBS1, SPINK1, CPD, MSN, LRP11, C7, and CLSTN3. For infarcts, key selected proteins included GSN, INHBA, and PPT1 (actin cytoskeleton), SCRG1 and PRSS23 (extracellular matrix), NLGN3, ROBO4, VWF, C7, PCDH17, FGFR1, NAGLU, ALAD, IGFBP4, NDRG2, NEU1, and FAM20C. The final model, trained on 75% of the data using the selected proteins, achieved median area under the receiver operating characteristic curve (AUC) values after 100 permuted runs of 0.715 for microbleeds (95% confidence interval: 0.678–0.753), 0.808 for infarcts (95% confidence interval: 0.794–0.849), 0.904 for white matter hyperintensities (95% confidence interval: 0.847–0.960), and 0.921 for PET AV45 (95% confidence interval: 0.885–0.957) when evaluated on 25% of the withheld data. (**Figures 4F-4I**). Notably, the top shared proteins across all three vascular outcomes mapped to ECM- and vasculature-associated modules (M7, M9, M10), significantly linked to CAA pathology (**Figure 4J** and **4K and Table S21)**. These results suggest that integrating proteomic data improves the prediction of neurovascular manifestations and highlights potential biomarkers for validation in independent cohorts.

### CSF protein biomarkers of vascular neuroimaging abnormalities map to CAA-enriched brain modules distinct from amyloid plaque pathology

Identifying CSF proteins associated with neuroimaging markers of vascular injury and cerebrovascular pathology is essential for distinguishing CAA-driven damage, paving the way for more accurate diagnoses. To achieve this, we systematically mapped CSF proteins associated with vascular imaging abnormalities to the cerebrovascular proteome network, revealing a molecular signature enriched in key brain modules linked to CAA (**Figure 1, Table S22**). Remarkably, we identified a cluster of three ECM brain modules: M7, M9, and M10 enriched with CSF proteins across all three vascular abnormalities underscoring their utility as highly specific discriminators of CAA and imaging manifestations (**Figure 5A**). Integration of PET AV45 with our vascular proteomic modules revealed six modules enriched in amyloid plaque-associated proteins, particularly those involved in synaptic and metabolic process, and ECM organization. Remarkably, CAA-enriched modules associated with neuroimaging abnormalities of vascular dysfunction (M7, M9, and M10) were largely independent of amyloid plaque burden via PET AV45 (**Figure 5B, Table S23**). In stark contrast, M37 exhibited no association with these vascular imaging abnormalities but overlapped with AV45-associated CSF proteins, reinforcing its distinct role in amyloid plaque pathology rather than vascular damage (**Figures 5A** and **5B**). This included the protein SMOC1 (*p value = 8.22×10^-56^, β = 0.24*), which showed the strongest association with PET AV45 signal (**Fig. 4E**), supporting prior studies that associated SMOC1 to amyloid PET signals^36,37^.

**Figure 5.**
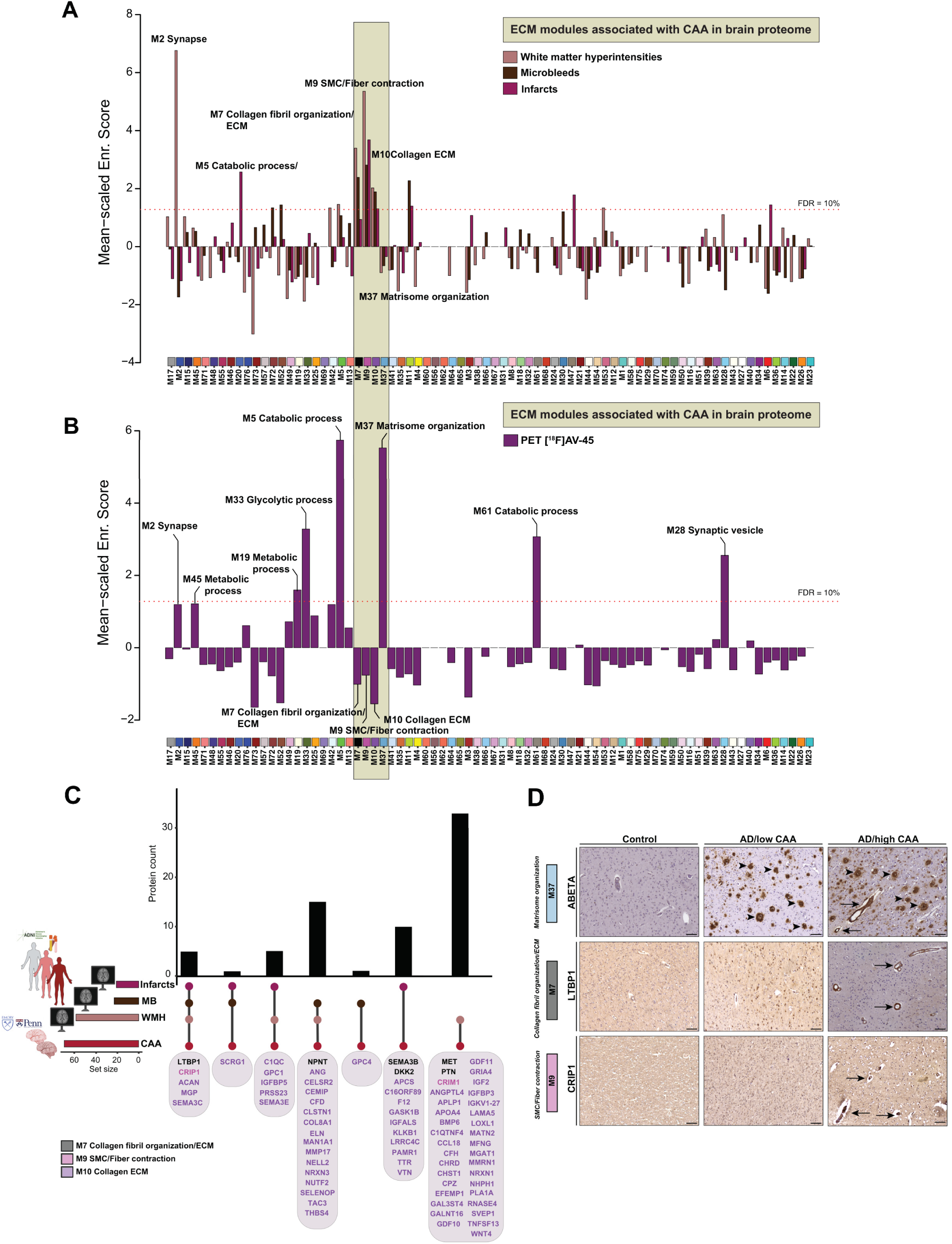
Cerebrovascular extracellular matrix (ECM) modules (M7, M9, and M10) link CSF biomarkers of vascular imaging abnormalities to CAA pathology. (A) CSF proteins associated with WMH, microbleeds, and infarcts were mapped to cerebrovascular protein network. The horizontal red dotted line indicates the threshold for permutation test false discovery rate (FDRs of 10%) above which the enrichment of proteins correlated with WMH, proteins was considered significant. Yellow box outlines modules: M7, M9, and M10 with significant enrichment of proteins linked to neuroimaging abnormalities. (B) Bar plot showing cerebrovascular modules enriched for CSF proteins associated with PET AV45. The horizontal red dotted line indicates the threshold for the permutation test (FDRs of 10%) above which the enrichment of PET AV45 associated proteins was considered significant. (C) The UpSet plot shows the overlap of proteins from modules M7, M9, and M10 that are associated with imaging vascular abnormalities (WMH, microbleeds, and infarcts) in the CSF proteome and CAA in the cerebrovascular proteome, highlighting the shared molecular signature across fluid- and tissue-based markers of neurovascular injury. (D) Immunohistological evaluation of expression patterns of ABETA, LTBP1 and CRIP1 in human postmortem cortical tissues from non-demented control, AD, AD with severe CAA. Arrows indicate the presence of these proteins in the cerebrovasculature, Scale bar, 100 μm.

To assess the generalizability of our findings, we leveraged a recently published CSF dataset from BioFINDER-2 study comprising of controls, MCI, and AD participants (total N=856)^38^. Similar to our study, CSF proteins, quantified using proximity extension assay (Olink®) technology (N = 2,943), were associated with vascular manifestations, including WMH, microbleeds, and infarcts, providing a detailed profile linked to cerebrovascular injury.^38^ (**Figure S9A, Table S24**). Using this Olink® proteomic dataset, we mapped dysregulated CSF proteins correlated to imaging abnormalities to our cerebrovascular network, once again identifying the ECM brain modules M7, M9, and M10 as significantly associated with vascular manifestations (**Figure S9B, Tables S25** and **S26**). Further analysis uncovered both unique and shared markers of the distinct vascular conditions (**Figure S9C**) and delineated the degree of concordance between the ADNI and BioFINDER-2 studies despite different proteomic platforms implemented (**Figure S9D**). Together, these findings reveal distinct, reproducible, and biologically meaningful CSF proteomic signatures associated with specific cerebrovascular lesions in AD, helping to disentangle their overlap with, and divergence from, canonical amyloid pathology.

### ECM proteins linked to vascular abnormalities show high specificity to CAA but not amyloid plaques

We prioritized validation of proteins from CAA-associated modules (M7, M9, and M10) based on their specific associations with vascular imaging abnormalities and their enrichment in isolated cerebrovascular fractions from AD brain. From this effort, we curated a panel of candidate proteins (**Table S27**) that demonstrated clear link to CAA in brain and MRI vascular abnormalities in CSF, revealing varying degrees of overlap across these phenotypes. Notably, a subset of proteins: LTBP1, CRIP1, MGP, ACAN, and SEMA3C emerged as significantly associated (*p value<0.05*) with all three vascular imaging abnormalities and CAA pathology, suggesting their central role as biomarkers of cerebrovascular defects in AD (**Figures 5C** and **5D** and **Figure 6**). To corroborate these findings at the tissue level, we employed immunohistochemistry (IHC) to assess the spatial distribution of key proteins in cortical brain tissue from individuals with confirmed severe CAA, low CAA, and non-AD controls. We observed pronounced levels of LTBP1 and CRIP1 in the cerebrovasculature of AD brain with high CAA burden, whereas these proteins were undetectable in AD cases with low CAA or in cognitively normal controls. Notably, LTBP1 and CRIP1 staining did not resemble pattern of parenchymal amyloid plaques, underscoring their specificity to the vascular compartment (**Figure 5D**). In contrast, staining with anti-Aβ antibody (M37 Matrisome member), that served as a reference, was broadly distributed in both vascular and parenchymal regions, indicating the presence of amyloid pathology (CAA and plaque) across both brain compartments (**Figure 5D**).

**Figure 6.**
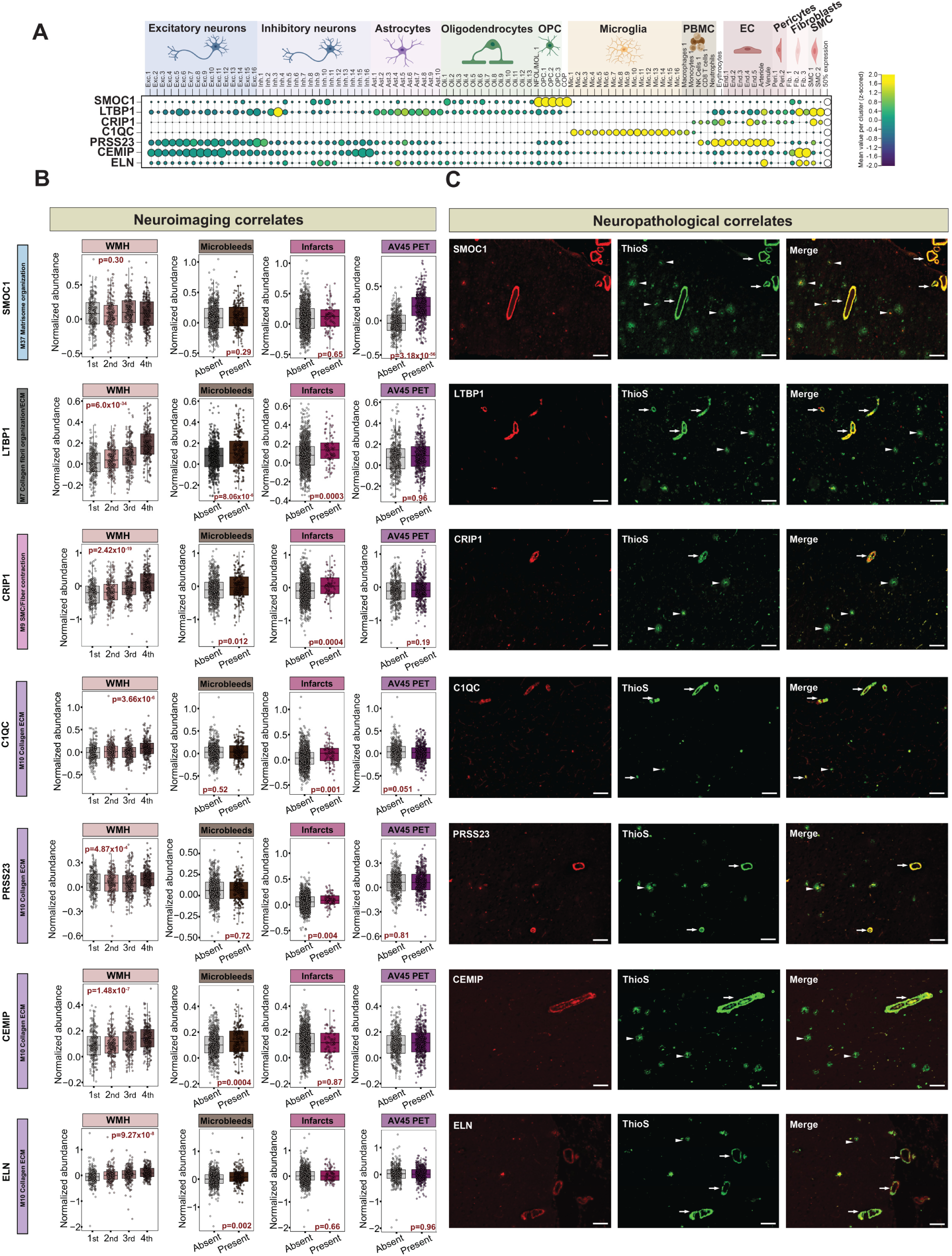
CSF and histological validation of vascular extracellular matrix (ECM) module proteins associated with neuroimaging biomarkers and amyloid pathology. (A) Single-nucleus RNA-sequencing data from prefrontal cortex of 437 ROSMAP individuals. The dot plot shows average gene expression for each cell type specific cluster and the fraction of nuclei within each cluster that show detection. The heat map illustrates mean z-scored value per cluster with yellow color indicating increase and purple showing decrease. NFOL/MOL, newly formed oligodendrocytes/mature oligodendrocytes; OPC, oligodendrocyte progenitor cells; COP, circulating osteogenic precursor. (B) Boxplots representing WMH, microbleeds, infarcts, and AV45 association with selected proteins. WMH burden was divided into four tertiles. Each subplot displays protein expression levels across groups defined by the presence or burden of the corresponding neuroimaging marker. Microbleeds, N=732 (194 positive and 538 negative); infarcts, N=1050 (81 positive and 969 negative), WMH, N=775; AV45, N=745 (384 positive, 361 negative). (C) Immunohistological evaluation of expression patterns of proteins from M7, M9, and M10 in human post-mortem cortical tissues from unimpaired control and AD with severe CAA. Thioflavin-S (thioS) stain was used to label fibrillar amyloid deposition. Arrows indicate the co-localization of these proteins with CAA, the arrowheads indicate their co-localization with amyloid plaques. Scale bar, 100 μm.

To gain insight into the cellular origins of selected proteins associated with CAA and correlated with vascular lesions, we analyzed single-nucleus RNA sequencing (snRNA-seq) data from 437 ROSMAP participants^39^. This analysis indicated that many of the CAA-associated proteins exhibit vascular-specific cell-type expression, localizing to fibroblasts and smooth muscle cells (e.g., LTBP1, CRIP1, CEMIP, ELN) as well as endothelial cells (e.g., CRIP1, ELN). In contrast, SMOC1 and C1QC were predominantly expressed in glial cell populations (**Figure 6A**). These findings suggest that CAA-associated proteins originate from distinct vascular and glial cell types highlighting their potential roles in vascular remodeling and integrity.

We next examined the spatial relationship between these proteins and amyloid pathology by co-staining adjacent brain sections with thioflavin-S (ThioS), a fluorescent dye that binds β-pleated sheet-rich amyloid aggregates^40,41^. ThioS staining enables the simultaneous visualization of parenchymal amyloid plaques and vascular amyloid deposits, allowing precise localization of amyloid pathology in tissue context. Consistently with previous findings^17,42^, SMOC1 was enriched in both vascular and parenchymal amyloid deposits, showing clear co-localization with ThioS-positive CAA and the dense cores of plaques, suggesting a broader amyloid affinity (**Figures 6B** and **6C**). Notably, LTBP1 and CRIP1, along with C1QC and PRSS23 (linked to WMH and infarcts) and CEMIP and ELN (associated with WMH and microbleeds), showed strong co-localization with ThioS-positive vascular amyloid deposits, but not with parenchymal plaques (**Figures 6B** and **6C**). This distinct spatial overlap between protein localization and vascular, rather than parenchymal amyloid highlights the specificity of these markers for cerebrovascular manifestations of AD. The concordance between CSF-based proteomic signatures of CAA and their histopathological validation highlights the robustness of our approach and reinforces the association between these proteomic changes and CAA-driven vascular pathology.

### CRIP1 is causally linked to AD and associated with WMH and blood pressure

To systematically connect inherited genetic risk to downstream molecular drivers of cerebrovascular pathology in AD, we leveraged our recent integration of AD GWAS results^43^ with CSF proteogenomic data from ADNI cohort using a proteome-wide association study (PWAS)^44^ (**Figures 1** and **7A**). This identified a genetically anchored panel of 24 CSF proteins associated with AD risk. Notably, 4 of those candidate risk proteins are ECM-related proteins, including LGALS3 and LRP2 (in module M7), CRIP1 (in module M9), and CTSH (in module M37). Since CRIP1 protein levels in CSF were positively associated with CAA pathology and all three vascular neuroimaging abnormalities (WMH, microbleeds, and infarcts) in ADNI, we next focused on the genetic variant linked to CSF CRIP1 variation, the protein quantitative trait locus (pQTL) rs35590487 (**Figure 7B**). We found that individuals carrying the minor allele of this variant displayed markedly reduced CRIP1 protein levels in CSF (N= 988, *p value = 1.79×10^-9^*) and less WMH (N=996, *p value = 0.03*), respectively (**Figures 7C** and **7D**). A significant reduction in WMH burden in rs35590487 minor allele (T) carriers was also observed even following adjustment for age (*p value=0.0173*), sex (*p value=0.0369*), and *APOE4* genotype (*p value=0.0216*). As WMH is a hallmark of small vessel disease and CAA, these findings not only implicate CRIP1 in genetically mediated cerebrovascular injury but also suggest the presence of a protective, allele-specific mechanism. Moreover, higher CRIP1 protein level in CSF correlated with higher systolic blood pressure (N=1087, *p value=1,32×10^-5^)* (**Figure 7E**), and carriers of the major allele of the CRIP1 rs35590487 variant showed a trend toward higher systolic blood pressure, although this did not reach statistical significance. (N=1293, *p value=0.091)* (**Figure 7F**). Together, these findings link CRIP1 to vascular homeostasis and suggest a broader role in cerebrovascular regulation.

**Figure 7.**
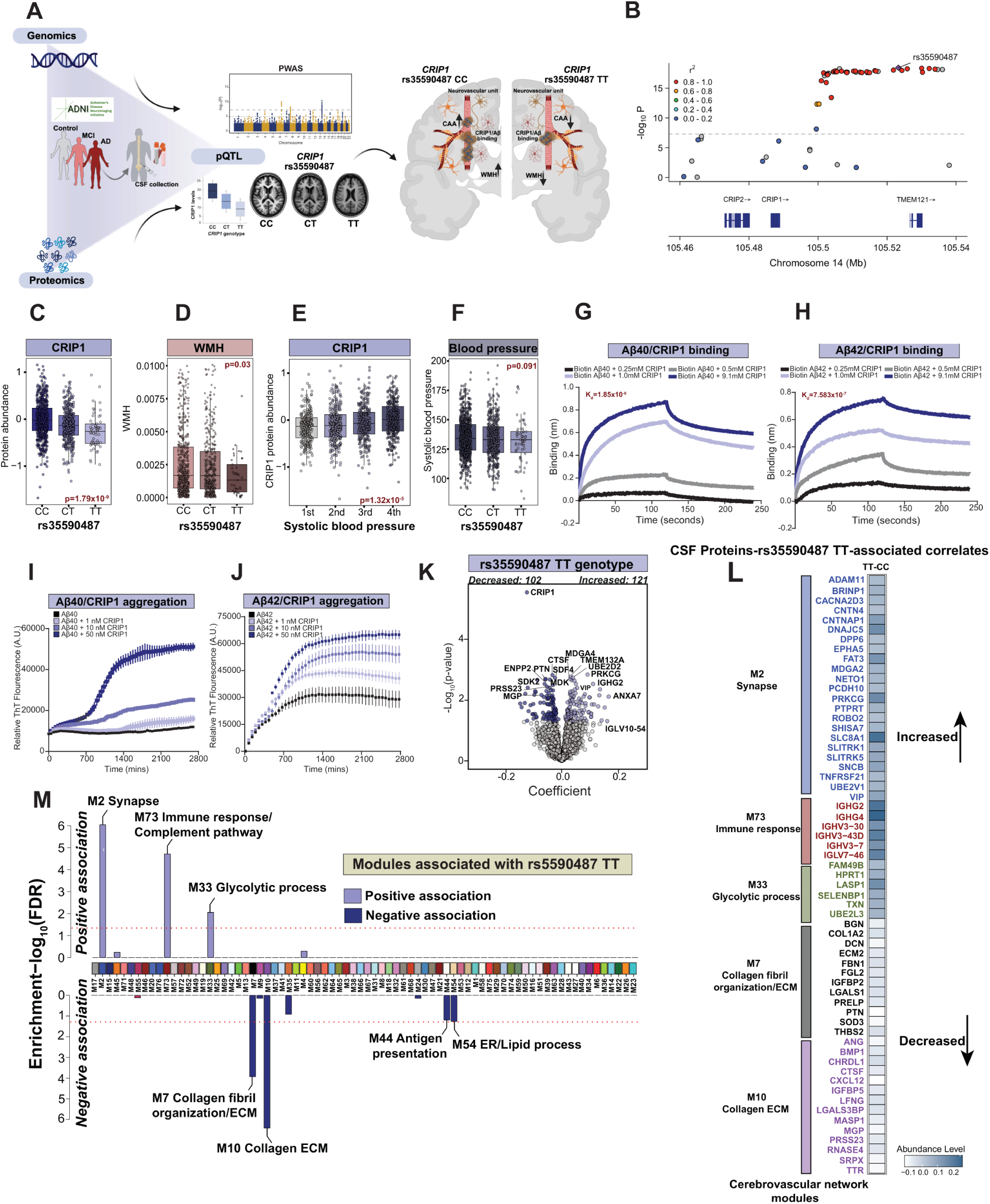
PWAS identifies CRIP1 as a causal protein linking AD risk to CAA and cerebrovascular dysfunction. (A) Schematic representation of the experimental workflow. The integration of AD GWAS results with ADNI CSF proteogenomic data via PWAS and Mendelian randomization led to the identification of CRIP1 as a candidate causal protein in AD. We also identified rs35590487 as the protein quantitative locus (pQTL) that influences the expression of CRIP1 in CSF. A hypothetical model of CRIP1 role in CAA formation. The major C allele (risk allele) of rs35590487 leads to more CRIP1 that binds amyloid in the vasculature impacting the severity of CAA and WMH. The minor T allele of rs35590487 is associated with lower levels of CRIP1 and less severe CAA occurrence in the blood vessels functioning as a protective factor. (B) LocusZoom plot of CRIP1 pQTL (within 100kb window). rs35590487 is the index pQTL for CRIP1 in the CSF summary-based Mendelian randomization for AD. (C) CRIP1 CSF pQTL - rs35590487 - was associated with the level of CRIP1 protein in CSF. The level of CRIP1 change in the allele dose-dependent manner in 988 ADNI participants (CC=558, CT=374, TT=56). (D) The major C allele of rs35590487 is associated with higher occurrence of WMH, whereas the minor allele of rs35590487 (T) was associated with lower occurrence of WMH in 996 ADNI participants (CC=581, CT=374, TT=41). (E) Higher CSF CRIP1 protein level was associated with higher systolic blood pressure in 1087 ADNI participants. (F) The minor allele of rs35590487 (T) has a trend toward lower systolic blood pressure in 1293 ADNI participants (CC=754, CT=476, TT=63). (G-H) The BLITZ assay showing binding of biotinylated Aβ40 (G) or Aβ42 (H) to increasing concentrations of recombinant human CRIP1 during 120 seconds, following their dissociation. The global dissociation constant (K_d_) was calculated for Aβ40 and Aβ42. (I-J) The thioflavine T (ThT) aggregation assay of recombinant human Aβ1-40 (I) or Aβ1-42 (J) with or without recombinant human CRIP1. (K) Volcano plots displaying differential abundance of CSF proteins at p<0.05 correlated with rs5590487 TT genotype after adjustment for age, sex, and diagnosis (Increased: 121, Decreased: 102) in ADNI participants (CC: N=558, TT=56). The x axis shows the regression coefficient, the y axis indicates −log 10 statistical p value for all proteins. Select proteins are labeled. (L) Heatmap exhibiting selected CSF proteins with positive and negative correlation to rs35590487 TT. Colors show a difference of abundance levels between CC and TT carriers. (M) Integrated approach of CSF proteomic dataset with cerebrovascular network. The horizontal red line indicates the threshold for permutation test false discovery rate (FDRs of 5%) above which the enrichment of CSF proteins correlated with rs5590487 TT genotype was considered significant. Significant modules are labeled.

To explore a functional link between CRIP1 and CAA, we assessed direct CRIP1– Aβ interactions using biolayer interferometry (BLI) and thioflavin-T aggregation assays. BLI assays were used to characterize the binding kinetics of recombinant CRIP1 to Aβ40 and Aβ42 peptides. For both peptides, CRIP1 exhibited a dose-dependent increase in binding signal during the association phase, indicating direct interaction (**Figures 7G** and **7H).** The dissociation phase revealed that CRIP1–Aβ40 complexes were less stable, with a faster signal decay, compared to CRIP1–Aβ42 complexes, which dissociated more slowly. The calculated dissociation constants (K_D_) were 1.85 × 10⁻^6^ M for Aβ40 and 7.583 × 10⁻⁷ M for Aβ42, confirming submicromolar affinity in both cases. In thioflavin-T aggregation assays, recombinant CRIP1 accelerated the nucleation and elongation phases of both Aβ40 (**Figure 7I**) and Aβ42 (**Figure 7J**). However, the addition of CRIP1, compared to Aβ42, induced more robust Aβ40 fibrillization, with increasing concentrations (**Figures 7I** and **7J**). These findings establish CRIP1 as both a genetically regulated and functionally active mediator of amyloid-associated vascular pathology in AD.

Finally, we evaluated the global impact of carrying two copies of the minor (T) allele of the *CRIP1* variant rs35590487 on the CSF proteome, compared to individuals homozygous for the major (C) allele, adjusting for age, sex, and clinical diagnosis. Homozygous TT carriers exhibited broad proteomic changes, including significant increases in synaptic- and metabolism-related proteins, and reductions in proteins such as MGP and PRSS23, both of which are directly associated with CAA pathology in brain (**Figure 6**, **Figures 7K** and **7L, Table S28**). We further integrated the differentially abundant proteins into our cerebrovascular proteomic network (**Figures 7L** and **7M** and **Table S29**). Proteins significantly elevated in TT compared to CC homozygote carriers were enriched in modules linked to synaptic function (M2), metabolism (M33), and immune regulation (M73). In contrast, proteins reduced in TT homozygote carriers were strongly enriched in ECM modules (M7, M10), lipid processing (M54), and antigen presentation pathways (M44). Notably, key ECM proteins from M7 and M10, many of which are associated with CAA and vascular lesions (**Figures 5** and **6**), were significantly lower in TT homozygotes (**Figures 7L** and **7M**). Overall, these results highlight a protective effect of the CRIP1 minor allele, potentially through enhanced synaptic and metabolic resilience and reduced CSF levels of proteins linked to CAA pathology and vascular abnormalities.

## Discussion

In this study, we implemented a comprehensive, multi-dimensional approach that integrates proteomic data from CSF and brain tissue with clinical, genetic, neuroimaging, and neuropathological assessments. This integrative framework enabled a compartment-specific exploration of disease signatures from the cerebrovasculature enhancing our ability to resolve CAA-specific molecular pathways that would likely be missed if brain and CSF data were analyzed independently. Through this approach, we identified ECM dysfunction as a prominent molecular feature of CAA, with four distinct ECM modules showing strong and reproducible associations with CAA pathological severity. Previous studies have consistently highlighted widespread ECM alterations in AD, reproducibly captured across both bulk brain tissue and vascular-enriched proteomic networks in multiple independent cohorts^12,17,42^. Notably, key ECM-associated proteins such as SMOC1, MDK, and SFRP1 are recurrently elevated and exhibit strong specificity to amyloid pathology, spanning both parenchymal plaques and vascular amyloid in CAA. By integrating our vascular proteomics with spatially resolved proteomic datasets^16^, we refined the molecular architecture of ECM disruption, revealing that ECM dysregulation in AD is not uniform, but instead follows compartment-specific patterns defined by distinct network modules. This distinction was further reinforced by PET AV45 imaging, which showed that ECM matrisome module M37 was strongly aligned with parenchymal amyloid but only weakly associated with CAA-related vascular pathology. Together, these findings underscore that vascular and parenchymal compartment in AD are governed by distinct ECM remodeling mechanisms and suggest that disentangling these compartmentalized processes will be essential for developing targeted interventions that preserve vascular integrity without inadvertently exacerbating parenchymal pathology.

Central to our findings was an unbiased, discovery-driven CSF proteomic approach that proved highly effective for identifying biomarkers of cerebrovascular pathology. We integrated CSF proteomic profiles from the ADNI cohort, specifically linked to vascular imaging abnormalities such as WMH, cerebral infarcts, and microbleeds, into our cerebrovascular proteomic network. This integration enabled the differentiation of ECM-enriched modules (M7, M9, and M10) that were selectively associated with CAA and other vascular pathologies, while remaining distinct from proteomic signatures related to parenchymal amyloid accumulation (M37). These CSF-based associations were independently validated in the deeply phenotyped BioFINDER-2 cohort using a different proteomic platform (Olink®), reinforcing the generalizability and translational potential of our candidate vascular biomarkers. Our findings align with a recent, large genome-wide association study (GWAS) that identified ECM genes as key contributors to WMH burden^45^, supporting the central role of matrisome pathways in cerebrovascular dysfunction and validating our proteomics-derived ECM modules as genetically linked markers in AD and small vessel disease.

Several top-ranked vascular proteins converged on pathways related to structural integrity, immune activation, and cellular plasticity within the vascular niche. LTBP1, a regulator of TGF-β signaling and known HTRA1 substrate, emerged as a strong candidate biomarker given its role in angiogenesis and its links to CADASIL^46,47^. Although LTBP2, a regulator of ECM integrity^48^, did not meet the significance threshold for CAA in brain tissue, its associations with WMH, microbleeds, and infarcts highlight its potential relevance to small vessel disease. Other notable markers (e.g., C1QC, PRSS23) were linked to WMH and infarcts, with C1QC involved in inflammation and vascular remodeling^49^ and PRSS23 potentially regulating endothelial-to-mesenchymal transition^50^. Finally biomarkers shared across WMH, microbleeds, and CAA, such as CEMIP and ELN underscore roles in matrix remodeling, vascular plasticity, and vessel stiffness^51,52,53,54^, further supporting a convergent molecular signature of small vessel disease. Beyond passive indicators of disease, these proteins represent mechanistically grounded targets with direct relevance for disease stratification and the redefinition of vascular contributions to cognitive decline.

While pQTL studies have become a foundational tool for linking genetic variants to protein abundance^55,56^, they rarely extend beyond statistical association to demonstrate causal mechanisms or functional consequences relevant to disease. To bridge this gap, we not only identified CRIP1 as a genetically regulated protein linked to vascular pathology in AD, but also demonstrated its mechanistic role, transforming it from a statistical association into a biologically validated disease effector. This functional validation further supports its potential causal role in AD. CRIP1 initially prioritized as a candidate causal AD gene through PWAS^44^, emerged as one of the few proteins consistently associated with multiple cerebrovascular imaging phenotypes in CSF and CAA pathology in brain. Importantly, a cis-pQTL (rs5590487) regulates CRIP1 levels in CSF, with the minor allele associated with both reduced CRIP1 expression and lower WMH burden, a rare example of genetically anchored protection against small vessel disease. This genetic signal was complemented by an observed association between elevated CRIP1 levels and higher systolic blood pressure, an established risk factor for small vessel disease^57^, thereby positioning CRIP1 at the intersection of AD genetic risk, vascular injury, and systemic hypertension^58,59^. The cysteine-rich intestinal protein (CRIP) family plays an important role in epithelial-mesenchymal transition, cell death, and immunity^60^. Multiple studies have linked CRIP1 expression to hypertension and stroke risk possibly by regulating interactions with cytoskeletal proteins leading to altered monocyte migration and invasion^58,59,61^. Importantly, our findings at the protein level support transcriptomic studies that have previously linked cumulative exposure to elevated systolic blood pressure, particularly in later life, with increased WMH volume^62,63^. In turn, anti-hypertensive treatment has been shown to attenuate WMH progression, supporting intensive blood pressure control as a strategy to preserve cerebrovascular integrity and prevent cognitive decline^64,65^.

Our findings raise the possibility that reducing CRIP1 expression, mimicking the protective effect observed in rs35590487-TT carriers, may offer a novel therapeutic approach for mitigating cerebrovascular damage and slowing progression of amyloid-associated dementias. This was further supported by widespread shifts in the CSF proteome, including increased levels of synaptic and metabolic proteins and decreased levels of ECM and CAA-associated proteins in individuals carrying two copies of the CRIP1 minor (T) allele at rs35590487. Crucially, we also provided direct functional evidence that CRIP1 binds both Aβ40 and Aβ42, accelerating their aggregation *in vitro*, and co-localizes with vascular amyloid deposits in postmortem brain tissue. These findings support a model by which elevated CRIP1 actively promotes amyloid nucleation and aggregation within vessel walls, likely contributing to increased WMH, infarcts, and microbleeds observed by MRI, while reduced CRIP1 levels may confer resilience to such injury (**Figure 7A**). Given the established role of CAA in driving ARIA^5,66^, these findings also raise the possibility that individuals with the CRIP1 CC genotype may be more susceptible to ARIA when treated with anti-Aβ monoclonal antibodies, while TT carriers may be relatively protected. Thus, by integrating genetic, imaging, pathological, and functional data, our study demonstrates that CRIP1 is a genetically regulated protein that links vascular dysfunction to amyloid pathology in AD.

## Conclusions

Clinically, these findings enhance the prioritization of protein biomarkers that reflect CAA pathology in the brain, offering a critical step toward distinguishing vascular amyloid involvement from parenchymal plaque pathology. The ability to detect cerebrovascular dysfunction through CSF biomarkers provides a non-invasive and more accurate diagnosis, which is especially valuable in the context of anti-Aβ immunotherapies, where patients with underlying CAA are at increased risk of ARIA, including cerebral edema and hemorrhage. By enabling stratification of patients based on underlying vascular pathology, these biomarkers can inform clinical decision-making, improve risk assessment, and guide the selection or monitoring of therapeutic interventions. Furthermore, the identification of both shared and distinct protein signatures linked to neuroimaging and neuropathology establishes a molecular framework for developing targeted, disease-modifying treatments that address the vascular component of cognitive impairment, an aspect that is increasingly recognized as essential to managing AD more effectively.

### Limitations of the study

Despite the strengths of this multi-dimensional, proteomics-driven framework and uncovering CRIP1 as a putative effector of CAA-related vascular dysfunction, some aspects of its biological function remain to be clarified. Future studies using CRIP1-targeted perturbation in vascular-specific models could help clarify its role in vascular biology and determine whether CRIP1’s association with CAA reflects a direct contribution to pathology or a broader adaptive or regulatory response. While such functional work is not necessary to support CRIP1’s utility as a biomarker, it could deepen our understanding of its therapeutic potential and shed light on the mechanisms by which genetic variation influences cerebrovascular outcomes. In addition to CRIP1, two candidate AD causal targets identified by PWAS from ADNI CSF, galectin-3 (LGALS3) and low-density lipoprotein receptor-related protein 2 (LRP2), also mapped to CAA-enriched vascular modules in the brain. LRP2 (i.e., megalin) directly interacts with clusterin (CLU), functioning as an endocytic receptor that facilitates the clearance of CLU-bound proteins such as Aβ^67,68^. Galectin-3 plays a critical role in microglial activation in response to amyloid pathology and has been implicated in modulating Aβ oligomerization and associated toxicity^69,70,71^. While neither protein was the primary focus of the current study, their genetic association with AD and biological relevance to amyloid and vascular processes highlight them as promising targets for future investigation. Finally, while this study identified CSF protein associations relevant to CAA-related vascular lesions, longitudinal analyses are essential to determine whether these biomarkers reflect disease trajectory and if changes in protein levels correspond to early events or late-stage pathology. Comparative studies across CSF and plasma may also provide key insights into the systemic detectability and translational potential of CSF-derived cerebrovascular biomarkers.

## Methods

### Cerebrovascular proteomics

#### Patient cohort characteristics-Emory and UPenn cohorts

Postmortem frozen frontal cortical tissues were obtained from the Emory University Brain Bank and the University of Pennsylvania (UPenn) Brain Bank using appropriate de-identification and under proper institutional review board protocols. Cases were selected based on the neuropathological assessment of Alzheimer’s disease (AD) or control at the time of autopsy. Specimens were banked at −80 °C after allocation by qualified pathologists according to institutional guidelines. The Emory and UPenn cohorts’ characteristics are presented in Table S1.

#### Cerebrovascular isolation

Frozen brain tissue (300 mg) was homogenized in ice-cold HBSS buffer with 0.1%BSA and 1% dextran (from *Leuconostoc mesenteroides*, Sigma D1537) in a glass Dounce homogenizer, as previously described^17^. An equal volume of 31% dextran was added to the homogenate and centrifuged at 8000 x g at 4C for 30 minutes leading to separation of myelin/parenchymal material (top layer) from vasculature (pellet). After the initial separation, the vascular pellet was suspended with 1 mL of 31% dextran buffer followed by the second step of centrifugation (at 4C for 30 minutes). After a series of washes, the vascular-enriched pellet was resuspended in HBSS with 0.1% BSA and the vessels were captured on 40um cell strainer and then collected in a new 50 mL falcon tube. Finally, the vessels were centrifuged followed by washes with ice-cold PBS. The vessel-enriched pellet was then lysed in 8M urea lysis buffer (8M urea, 10mM Tris, 100mM NaH2PO4, pH 8.5) with HALT protease and phosphatase inhibitor cocktail (Thermo Fisher Scientific).

#### Tissue Homogenization and Protein Digestion

Samples were homogenized in 8 M urea lysis buffer (8 M urea, 10 mM Tris, 100 mM NaH2PO4, pH 8.5) with HALT protease and phosphatase inhibitor cocktail (ThermoFisher) using a Bullet Blender (NextAdvance). Each Rino sample tube (NextAdvance) was supplemented with ∼100 μL of stainless-steel beads (0.9 to 2.0 mm blend, NextAdvance) and 300 μL of lysis buffer. Tissues were added immediately after excision and homogenized with bullet blender at 4 °C with 2 full 5 min cycles. The lysates were transferred to new Eppendorf Lobind tubes and sonicated for 3 cycles consisting of 5 s of active sonication at 30% amplitude, followed by 15 s on ice. Samples were then centrifuged for 5 min at 15,000 x g and the supernatant transferred to a new tube. Protein concentration was determined by bicinchoninic acid (BCA) assay (Pierce). For protein digestion, 100 μg of each sample was aliquoted and volumes normalized with additional lysis buffer. Samples were reduced with 5 mM dithiothreitol (DTT) at room temperature for 30 min, followed by 10 mM iodoacetamide (IAA) alkylation in the dark for another 30 min. Lysyl endopeptidase (Wako) at 1:25 (w/w) was added, and digestion allowed to proceed overnight. Samples were then 7-fold diluted with 50 mM ammonium bicarbonate. Trypsin (Promega) was then added at 1:25 (w/w) and digestion proceeded overnight. The peptide solutions were acidified to a final concentration of 1% (vol/vol) formic acid (FA) and 0.1% (vol/vol) trifluoroacetic acid (TFA) and desalted with a 30 mg HLB column (Oasis). Each HLB column was first rinsed with 1 mL of methanol, washed with 1 mL 50% (vol/vol) acetonitrile (ACN), and equilibrated with 2×1 mL 0.1% (vol/vol) TFA. The samples were then loaded onto the column and washed with 2×1 mL 0.1% (vol/vol) TFA. Elution was performed with 2 volumes of 0.5 mL 50% (vol/vol) ACN.

#### Isobaric Tandem Mass Tag (TMT) Peptide Labeling

Each sample (containing 20 μg of peptides) was re-suspended in 100 mM TEAB buffer (100 μL). The TMT labeling reagents (5mg; TMTpro^TM^ 16-plex Label Reagent, Lot: XC342533, ThermoFisher Scientific) were equilibrated to room temperature, and anhydrous ACN (256 μL) was added to each reagent channel. Seven N channels were used for labeling (127N to 133N). Each channel was gently vortexed for 5 min, and then 41 μL from each TMT channel was transferred to the peptide solutions and allowed to incubate for 1 h at room temperature. The reaction was quenched with 5% (vol/vol) hydroxylamine (8 μl) (Pierce). All channels were then combined and dried by SpeedVac (LabConco) to approximately 150 μL and diluted with 1 mL of 0.1% (vol/vol) TFA, then acidified to a final concentration of 1% (vol/vol) FA and 0.1% (vol/vol) TFA. Labeled peptides were desalted with a 200 mg C18 Sep-Pak column (Waters). Each Sep-Pak column was activated with 3 mL of methanol, washed with 3 mL of 50% (vol/vol) ACN, and equilibrated with 2×3 mL of 0.1% TFA. The samples were then loaded and each column was washed with 2×3 mL 0.1% (vol/vol) TFA, followed by 2 mL of 1% (vol/vol) FA. Elution was performed with 2 volumes of 1.5 mL 50% (vol/vol) ACN. The eluates were then dried to completeness using a SpeedVac.

#### High-pH Off-line Fractionation

Dried samples were re-suspended in high pH loading buffer (0.07% vol/vol NH4OH, 0.045% vol/vol FA, 2% vol/vol ACN) and loaded onto a Water’s BEH 1.7 um 2.1mm by 150mm. An Thermo Vanquish was used to carry out the fractionation. Solvent A consisted of 0.0175% (vol/vol) NH4OH, 0.01125% (vol/vol) FA, and 2% (vol/vol) ACN; solvent B consisted of 0.0175% (vol/vol) NH4OH, 0.01125% (vol/vol) FA, and 90% (vol/vol) ACN. The sample elution was performed over a 25 min gradient with a flow rate of 0.6 mL/min. A total of 192 individual equal volume fractions were collected across the gradient and subsequently pooled by concatenation into 96 fractions and dried to completeness using a SpeedVac.

#### Liquid Chromatography Mass Spectrometry

All fractions were resuspended in an equal volume of loading buffer (0.1% FA, 0.03% TFA, 1% ACN) and analyzed by liquid chromatography coupled to tandem mass spectrometry. Peptide eluents were separated on custom made fused silica column (15 cm × 75 μM internal diameter (ID) packed with Dr. Maisch 1.5um C18 resin) by a Vanquish Neo (ThermoFisher Scientific). Buffer A was water with 0.1% (vol/vol) formic acid, and buffer B was 80% (vol/vol) acetonitrile in water with 0.1% (vol/vol) formic acid. Elution was performed over a 15 min gradient. The gradient was from 1% to 99% solvent B. Peptides were monitored on a Orbitrap Astral mass spectrometer (ThermoFisher Scientific) fitted with a high-field asymmetric waveform ion mobility spectrometry (FAIMS Pro) ion mobility source (ThermoFisher Scientific). One compensation voltages (CV) of - 45 was chosen for the FAIMS. Each cycle consisted of one full scan (MS1) was performed with an m/z range of 400-1500 at 120,000 resolution at standard settings and as many tandem (MS/MS) scans in 1 second. The higher energy collision-induced dissociation (HCD) tandem scans were collected at 35% collision energy with an isolation of 0.7 m/z and a maximum injection time set to 20ms. Dynamic exclusion was set to exclude previously sequenced peaks for 20 seconds within a 5-ppm isolation window.

#### Immunohistological evaluation

To validate the expression patterns of proteins of interest, human postmortem brain tissues were obtained from the Emory University Brain Bank from age-matched individuals diagnosed with AD with high CAA, AD with low CAA, and non-demented controls. For histological analyses, paraffin-embedded frontal cortical sections were cut at 5 μm, then deparaffnized using xylene and rehydrated in a graded series of alcohols. Antigen retrieval was achieved by incubating tissue sections in distilled water for 30 min under high temperature. The brain sections were incubated with 3% hydrogen peroxidase for 5 min and blocked in goat serum for 1 hr, followed by overnight incubation with anti-Abeta, anti-LTBP1 (Invitrogen, PA5-45075, 1:250), anti-CRIP1 (Invitrogen PA5-24643, 1:50). After washes, the sections were incubated with biotinylated antibody for 30 min, followed by incubation with ABC reagent (Vectastain ABC-HRP kit, Vector Laboratories). The sections were then dehydrated, counterstained with hematoxylin, and coverslipped using permanent mounting media. The imaging was performed using Keyence microscope.

To assess the co-localization of target proteins with CAA or amyloid plaques, paraffin-embedded frontal cortical sections were processed through deparaffinization, rehydration, antigen retrieval, and blocking as described above. Sections were then incubated with the following primary antibodies: anti-LTBP1 (Invitrogen, PA5-45075, 1:250), anti-CRIP1 (PA5-24643, 1:50), anti-C1QC (PA5-35084, 1:25), anti-PRSS23 (PA5-56516, 1:100), anti-CEMIP (PA5-106288, 1:100), and anti-ELN (PA5-76676, 1:50).

After primary incubation, sections were treated with Cy3-conjugated secondary antibodies for 2 hours at RT. Following washes, sections were stained with 1% thioflavin S for 10 minutes at RT and counterstained with Sudan black B. Imaging was conducted using a Keyence microscope.

The expression profiles of proteins of interest was assessed using snRNA-seq data from the ROSMAP cohort (https://vmenon.shinyapps.io/rosmap_snrnaseq/).

#### Data analysis Database search

767 raw files (across 8 TMT-18 plexes, rawfile b03_09 was excluded) were searched using FragPipe (version 21.1). The FragPipe pipeline relies on MSFragger (version 4.0)^72,73^ Percolator^74^ for peptide identification, MSBooster^75^ and Percolator^74^ for false discovery rate (FDR) filtering and downstream processing. The search was performed with a database of canonical Human proteins downloaded from Uniprot (20,402; accessed 02/11/2019), as well as sequences for specific peptides for APOE4 and 2 alleles^23^ alongside ABETA40 and 42 peptides (total of 20405 sequences). The workflow used in FragPipe followed default TMT-16 plex parameters, used for both TMT-16 and TMT-18 experimental design. Briefly, precursor mass tolerance was −20 to 20 ppm, fragment mass tolerance of 20 ppm, mass calibration and parameter optimization were selected, and isotope error was set to −1/0/1/2/3. Enzyme specificity was set to fully-tryptic and up to two missing trypsin cleavages were allowed. Peptide length was allowed to range from 7 to 50 and peptide mass from either 200 to 5,000 Da. Variable modifications that were allowed in our search included: oxidation on methionine, N-terminal acetylation on protein, N-terminal acetylation on peptide along with off-target TMT tag modification on Serine, Threonine and Histidine, with a maximum of 3 variable modifications per peptide. The FDR threshold was set to 1% and protein and peptide abundances were quantified using Philosopher for downstream analysis. Before performing any abundance analysis, the data from all batches were merged and protein levels were first scaled by dividing each protein intensity by intensity sum of all proteins in each sample followed by multiplying by the maximum protein intensity sum across all nine samples. Instances where the intensity was ‘0’ were treated as ‘missing values’.

#### Batch correction of proteomics

Eight batches of TMT 18-plex each with one global internal standard (GIS, all sample mixture), and the fourth batch with a second GIS sample, were initially corrected for batch using TAMPOR, an algorithm we developed to leverage the ratio of samples over GIS within and across batches, stabilizing and minimizing variance due to batch iteratively median-centering both proteins and samples within the two-dimensional proteomic abundance matrix, as previously described^76^. Using the option useAllNonGIS=FALSE the correction converged immediately, in two iterations. Code for TAMPOR as an R function is available from https://www.github.com/edammer/TAMPOR.

#### Quantification of cell-type composition in vascular-enriched brain-derived proteomes

To estimate the cellular provenance of the log₂ proteomic abundance profiles generated in the present study we applied ensemble deconvolution as implemented in the *EnsDeconv* R package (v1.2-0)^77^. The package includes a curated list of human cortical single-cell reference matrices, which were used without modification and constitute the prior expression signatures against which all proteomic samples were deconvolved. Proteins were mapped from HGNC gene symbol to Ensembl gene IDs (Ensembl 104 / GRCh38 December 2021 archive) through the biomaRt R package. When several proteins mapped to the same gene, the row with the largest variance across samples was retained using the collapseRows() function of *WGCNA*. The ensemble method of the package’s EnsDeconv() function leveraged CIBERSORT, FARDEEP, NNLS, MUSIC, DWLS, SCDC and other algorithms with all possible deconvolution methods used (params: dmeths=NULL). Run time with 37 parallel process threads was 11 minutes. Common settings also included 50 markers selected from each reference cell type profile, Marker.Method=”t” (two-sample *t* statistic), and no additional normalization of the input data. Returned relative cell-type proportions were examined by heat-map (pheatmap v1.0.12), confirming coherent enrichment patterns across vascular, glial and neuronal classes (**Figure S1**).

The 118 non-GIS sample proteomic abundances were subjected to nonparametric bootstrap regression with a model including endothelial proportion, age, PMI, and TMT batch. Each modeled trait value times the median estimated coefficient of variation from 1000 iterations of fitting for each protein was subtracted from the log_2_(abundance) matrix. VariancePartition violin plots were generated to gauge the success of each data cleanup step, by visualizing the percetn variance explained by traits—from the starting data to the abundance matrix following TAMPOR, to that following regression.

#### WGCNA definition of vascular network modules

A weighted protein co-expression network for the regressed cerebrovasculature log_2_(abundance) matrix of 118 samples with n=11,408 proteins having less than 50 percent missing values was built using R v4.2.3 WGCNA v1.72-1 blockwiseModules() function with the following settings. Power was 8, deepSplit was 3, minimum module size was 10, merge cut height was 0.07, TOMDenom “mean,” bicor correlation for adjacency/TOM calculation, a signed network type, partitioning about medioids staging (pamStage), pamRespectsDendro, a reassignment threshold p value of 0.05, and a maximum block size larger than the total number of proteins, i.e., built in a single block. Following blockwiseModules definition of the 76 modules, the table of kMEs calculated using bicor, and module membership based on this was iteratively checked to reassign module members subject to these rules: (1) maximum difference from the highest kME of the module to which a protein could be assigned is 0.10 (2) module membership must be assigned if any kME is greater than 0.30, and (3) must be unassigned (grey) if all kMEs are less than 0.30. The rules were met within 5 iterations out of 30 possibly allowed.

#### Statistics, enriched ontology determination, and visualization

Statistics and graphical visualization were performed in R v4.2.2. Volcano plots used the custom plotvolc function available from https://www.github.com/edammer/parANOVA, which also provides the parANOVA.dex function for fast parallel calculation of one-way ANOVA+Tukey-corrected pairwise comparison p values, with fallback to Bonferroni correction for Tukey p values < 1e-8.5. One-tailed Fisher’s exact test heatmaps for enrichment of gene products in modules were rendered using https://www.github.com/edammer/CellTypeFET. Ontologies enriched by module were determined using GOparallel function and supporting online resources which are available via https://www.github.com/edammer/Goparallel. Module iGraph was rendered using the buildIGraphs function available from https://www.github.com/edammer/netOps. Additional visualization layout and formatting were performed in Illustrator 2025. All schematic illustrations were made using BioRender.

### Bulk brain proteomics

#### Patient cohort characteristics of Rush ADRC and-ROSMAP cohort

Phenotype data were collected from community-dwelling older adults enrolled in one of five ongoing cohort studies of aging conducted by the Rush Alzheimer’s Disease Center: the Religious Orders Study (ROS) and the Rush Memory and Aging Project (MAP)^78^, the Minority Aging Research Study (MARS)^79^, the Rush Alzheimer’s Disease Center African American Clinical Core (AA Core)^79^, and the Rush Alzheimer’s Disease Center Latino Core (Latino Core)^80^. Participants were enrolled without known dementia who consent to annual clinical evaluations and brain donation upon death. ROS, initiated in 1994, includes nuns, priests, and brothers residing across the United States. MAP, which began in 1997, includes participants living in retirement communities, private residences, or subsidized housing in northeastern Illinois. MARS and AA Core, which began in 2004 and 2008 respectively, recruit older African Americans, and the Latino Core, which began in 2015, recruit older Latino adults from the same geographical region as MAP. All studies follow harmonized data acquisition protocols administered by a common research team, enabling integrated analyses. Further methodological details have been described previously. All study procedures were approved by the Institutional Review Board of Rush University Medical Center. Data are available for general research use according to the following requirements for data access and data attribution (https://adknowledgeportal.synapse.org/#/DataAccess/Instructions). ROSMAP resources can be requested at www.radc.rush.edu.

#### Neuropathological assessment of CAA and AD

Paraffin-embedded sections were prepared from four regions: midfrontal, midtemporal, angular, and calcarine cortices. Immunohistochemical methods using one of the three monoclonal antibodies against Aβ were used for scoring Aβ deposition in meningeal and parenchymal vessels. The three antibodies were 4G8 (1:9000; Covance Labs, Madison, WI), 6F/3D (1:50; Dako North America Inc., Carpinteria, CA), and 10D5 (1:600; Elan Pharmaceuticals, San Francisco, CA).

For scoring CAA, we followed a recently proposed protocol^81^. CAA severity in each of the four regions was scored using a semiquantitative scale indicating degree of Aβ deposition in each vessel and number of affected vessels. The maximum of the meningeal and parenchymal CAA scores from each region were averaged for computation of an ordinal 4-level CAA summary score (none, mild, moderate, severe). For descriptive purposes, we used a binary variable indicating presence of moderate-severe CAA.

Modified Bielschowsky silver stain was used for staining sections from 5 regions of midfrontal, midtemporal, inferior parietal, entorhinal cortices, and hippocampus and visualizing the AD pathological hallmark of neuritic plaques. Regional counts of neuritic plaques were divided by their standard deviations and averaged across brain regions to make composite scores for neuritic plaques. The stained sections were also reviewed for adjudicating an AD pathological diagnosis. In addition, immunohistochemistry quantifies the burden of PHF tau tangles from the 8 brain regions, as previous described^82^.

#### TMT-MS, Database search, and data QC

TMT-MS–based proteomics on dorsolateral prefrontal cortex tissues from the Rush ADRC and ROSMAP cohorts have been reported previously^23,56,83^. Briefly, all cases (n=982 cases across 63 batches) were digested as described, labeled with TMTpro reagents, fractionated by high-pH reverse-phase chromatography, and analyzed by LC-MS/MS on an Orbitrap mass spectrometer. For round 1: 5mg; TMTproTM 16-plex Label Reagent Lot: #XC342532 (ThermoFisher Scientific), TMTpro-134C & TMTpro-135N Label Reagent, 5mg, Lot #: XB338618 (ThermoFisher Scientific) were used. For round 2: TMTpro^TM^ 16-plex Label Reagent Set, 1×5mg, Lot #: VH311511 was used. For round 3: TMTpro 16plex Label Reagent Set, 1x 5mg, Lot #: VH311511 (ThermoFisher Scientific) was used.

All raw files underwent a database search using Fragpipe (version 19.0) for DLPFC dataset. The database search parameters have been described elsewhere^23,84^. Initially, mzML files were created from the original MS.raw files for frontal (5374 raw files across 63 batches) using the ProteoWizard MSConvert tool (version 3.0) with specific options, including “Write index,” “TPP compatibility,” “Use zlib compression,” and a “peakPicking” filter setting. Following the creation of mzML files for each set, they were subjected to a search using MSFragger (version 3.5). The human proteome database used contained 20,402 sequences (Swiss-Prot, downloaded February 11, 2019) along with corresponding decoys and common contaminants. The search settings included a precursor mass tolerance of −20 to 20 ppm, a fragment mass tolerance of 20 ppm, mass calibration, parameter optimization, isotope error set to −1/0/1/2/3, strict-trypsin enzyme specificity, and allowance for up to two missed cleavages. Fully enzymatic cleavage type, peptide length (7 to 50), and peptide mass (200 to 5000 Da) criteria were defined. Variable modifications included oxidation on methionine, N-terminal acetylation on protein, and TMTpro modification on the peptide N-terminus, with a maximum of three variable modifications per peptide. Static modifications comprised isobaric TMTpro (TMT16) modifications on lysine, along with carbamidomethylation of cysteine. In the Post-MSFragger (version 3.6) search, Percolator was used for Peptide-Spectrum Match validation, succeeded by Philosopher (version 4.6.0) for protein inference using ProteinProphet and false discovery rate (FDR) filtering. Reports containing quantified peptides and UniprotID-identified proteins with FDR < 1% were generated. Following quantification, protein amounts were first scaled by dividing each protein intensity by all reporter ion intensities of the TMT channel (each sample) followed by multiplying the maximum channel specific protein intensity in the channel.

N=11349 proteins were quantified in N=982 samples. Only proteins with non-missing percentage >30% were retained. Log2ratio transformation in each sample was performed resulting in N=10030 proteins in N=982 samples. PCA analysis was performed to detect and remove sample outliers, leading to removal of 11 outliers (N=10030 proteins in N=971 samples). Regression analysis was performed to remove any variance due to postmortem interval and TMT batch, resulting in N=10030 proteins in N=971 samples^56^.

#### Statistical analyses APOE4 association

Similar to previously described methods^32^, linear regressions were used to examine associations of APOE ɛ4 with proteins (N=10,030) across N=873 individuals. All the linear regressions were controlled for age at death and sex. To correct for multiple testing, the p values of all the analyses were adjusted using Bonferroni method.

#### CAA association

Ordinal logistic regression models were used for examining the associations of the proteins with CAA in N=887 participants. Proteins (N=10,030) were tested in 2 sets of parallel models. The first set of models were adjusted for age at death, sex and neuritic plaque and the second set of models were adjusted for age at death, sex and PHF tau tangle density. P values were corrected by the Bonferroni method to address multiple testing inflation of type I error.

#### Module preservation in bulk proteome

Vascular protein network module preservation in ROSMAP bulk cortical proteome was performed using the R v4.2.3 WGCNA v1.72-1 module. Preservation function over 500 permutations with the 118-sample vascular protein abundance matrix and module assignments as template network, and the 971 ROSMAP sample bulk cortex protein abundance matrix as target.

#### Cerebrovascular module association to CAA or APOE4

To assess how cerebrovascular network modules relate to CAA after adjustment for neuritic plaques (10,015) and tau tangles (10,015) or APOE4 (10,017) we analyzed proteins identified by proteome-wide association study. Gene products along with corresponding p values were evaluated for enrichment within cerebrovascular network modules using a permutation test (10,000 permutations) in R. Exact p values were calculated using the ‘permp’ function from the statmod package. To quantify module-specific enrichment, we calculated Z scores by comparing the average p value of genes within each module to the distribution of mean p-values from 10,000 random permutations. The Z score was computed as the difference between the observed and permuted mean p-values, normalized by the standard deviation of the permuted distribution. This approach mirrors the enrichment analysis applied to MAGMA-derived gene-level statistics for genome-wide association study, as described in previous work^12^ (available at https://www.github.com/edammer/MAGMA.SPA).

### CSF proteomics

#### Patient cohort characteristics-ADNI cohort

Data used in the preparation of this article were obtained from the Alzheimer’s Disease Neuroimaging Initiative (ADNI) database (adni.loni.usc.edu). The ADNI was launched in 2003 as a public-private partnership, led by Principal Investigator Michael W. Weiner, MD. The primary goal of ADNI has been to test whether serial magnetic resonance imaging (MRI), positron emission tomography (PET), other biological markers, and clinical and neuropsychological assessment can be combined to measure the progression of mild cognitive impairment (MCI) and early Alzheimer’s disease (AD).

ADNI dataset comprises CSF collected at baseline visits which were assayed from 1,104 participants. ADNI is an ongoing longitudinal, multicenter observational study designed to discover imaging and biochemical biomarkers for diagnosing and tracking AD^85,86^. In particular, ADNI aims to evaluate whether MRI, PET, other biological markers, and clinical and neuropsychological assessments can be combined to track the progression of MCI and early-stage AD (http://adni.loni.usc.edu). The ADNI study was approved by the Institutional Review Boards at each participating ADNI site (see full list here: (http://adni.loni.usc.edu). All procedures were performed in accordance with relevant guidelines and regulations, and informed consent was obtained from all subjects prior to enrollment. Data used in the present study were downloaded on Oct 20th, 2024, and inclusion is thus restricted to subjects whose data was uploaded to the ADNI database prior to this date.

Participant recruitment for ADNI is approved by the Institutional Review Board of each participating site. All ADNI participants undergo standardized diagnostic assessment that renders a clinical diagnosis of either control, MCI, or AD using standard research criteria. Control participants had no subjective memory complaints, tested normally on Logical Memory II of Weschler Memory Scale, had an MMSE between 24–30, and a CDR of 0 with memory box score of 0. A subject is diagnosed as MCI if the study participant (i) reports concern due to impaired memory function; (ii) obtains a Mini Mental State Examination (MMSE) score between 24 and 30; (iii) a Clinical Dementia Rating Scale (CDR) score of 0.5; (iv) a score lower than expected (adjusted for years of education) on the Wechsler Memory Scale Logical Memory II (WMS-II); and (v) reports preserved function of daily living. AD participants also exhibited subjective memory concerns but also met NINCDS/ARDA criteria for probable AD. AD participants also showed abnormal memory function on Logical Memory II subscale from the Weschler Memory Scale, an MMSE of 20–26, and CDR of 0.5 or 0.1. Inclusion criteria for the current study were enrollment in ADNI1/GO or ADNI2/3, an available baseline CSF sample, and longitudinal cognitive outcome. ADNI traits can be requested at http://adni.loni.usc.edu.

#### Structural MRI processing

Each subject underwent a standardized 3T MR imaging protocol. Imaging sequences included T2* gradient recalled-echo, T1-weighted 3D MPRAGE, and FLAIR sequences (http://adni.loni.usc.edu/methods/documents/mri-protocols/). Before any subject was scanned using this protocol, an ADNI phantom was used to assess linear and nonlinear spatial distortion, signal-to-noise ratio, and image contrast, which were reviewed by a single quality-control center to ensure harmonization among the sites.

#### Microbleeds

Microbleeds were quantified visually by a board-certified neuroradiologist with subspecialty certification at Mayo Clinic on the T2* gradient recalled-echo sequences. Microbleeds were defined as hypointense lesions within the brain parenchyma that measured <10 mm on the gradient recalled-echo sequence. Only microbleeds that were considered definite were included in the analysis. Microbleeds were classified visually by location as 1) deep gray matter/infratentorial, if they involved the basal ganglia, thalami, brain stem, or cerebellum, or 2) lobar, if they involved other regions of the brain parenchyma, however, all microbleed types were grouped together for this study. More details could be found in earlier studies^87^ and on ADNI website. We categorized microbleeds as either 0 (absent) and ≥1 (present).

#### WMH

White matter hyperintensities (WMH) were quantified using a multi-step pipeline combining high-resolution 3D T1 and FLAIR MRI at the DeCarli Lab (UC-Davis). Preprocessing included non-brain tissue removal via an atlas-based method, co-registration of FLAIR to T1 using FLIRT, and inhomogeneity correction through bias field estimation with B-spline deformation^88,89^. Each subject’s images were non-linearly aligned to a common template space. WMH segmentation was performed using a modified Bayesian framework that incorporated histogram-based likelihood estimates, spatial priors from over 700 manually labeled cases, and tissue class constraints^90^. Voxels were labeled as WMH if their probability exceeded 3.5 standard deviations above the white matter mean. These WMH masks were then transformed back to native space for volume quantification. Final tissue segmentation (gray matter, white matter, CSF, and WMH) was achieved via an Expectation-Maximization algorithm refined with a Markov Random Field model to iteratively update Gaussian appearance models and spatial priors, enhancing segmentation accuracy^91,92^.

#### Infarcts

Infarcts assessment was performed in the DeCarli Lab (UC-Davis), using a manual visual assessment. The presence of MRI infarction is determined from the size, location and imaging characteristics of the lesion. Only lesions 3mm or larger qualified for consideration as cerebral infarcts. We categorized infarcts as either 0 (absent) and ≥1 (present).

#### Florbetapir A(V-45) Image Processing

This study includes amyloid (^18^F-AV45) PET scans obtained through the ADNI database. Preprocessed Florbetapir PET images from ADNI, with realigned frames, head position corrected through linear transformation, standardized voxel size, and smoothed to a uniform resolution of 6mm, were used in this study. Florbetapir PET scans were analyzed in each participant’s native space, using their structural MRIs acquired closest to the florbetapir PET scan. The structural MRIs were segmented into cortical regions of interest and reference regions for each subject using FreeSurfer. The florbetapir PET data were then realigned, and the mean of all frames was used to co-register the florbetapir data with the corresponding structural MRI. For each subject, cortical standardized uptake value ratio (SUVR) images were generated by dividing voxel-wise florbetapir uptake by the average uptake from the whole cerebellum reference region.

#### Cognitive Assessments

Cognitive assessments in ADNI include a comprehensive battery designed to evaluate multiple domains of cognitive function relevant to AD progression. Among these, the Alzheimer’s Disease Assessment Scale–Cognitive Subscale (ADAS-Cog) is a widely used clinical measure for assessing cognitive impairment. We focused on the 11-item version (ADAS-Cog11), which evaluates domains such as memory, language, orientation, and praxis. Scores range from 0 to 85, with higher scores reflecting greater impairment. The ADAS-Cog11 has been extensively validated and is commonly used as an outcome measure in clinical trials and observational studies to track cognitive decline.

#### Isobaric Tandem Mass Tag (TMT) Peptide Labeling

Each sample was re-suspended in 100 mM TEAB buffer (50 μL). The TMT labeling reagents (5mg; TMTpro^TM^ 16-plex Label Reagent, Lot: # YA357799; 134C and 135N lot# YB370079, ThermoFisher Scientific) were equilibrated to room temperature, and anhydrous ACN (200 μL) was added to each reagent channel. Each channel was gently vortexed for 5 min, and then 10 μL from each TMT channel was transferred to the peptide solutions and allowed to incubate for 1 h at room temperature. The reaction was quenched with 5% (vol/vol) hydroxylamine (5 μl) (Pierce). All channels were then combined and dried by SpeedVac (LabConco). The combined sample was then resuspended in 500 uL of 0.1% TFA and then diluted 1:1 with 4% H3PO4 and desalted with a 30 mg MCX column (Waters). Samples were loaded onto the MCX column and then washed with 100 mM ammonium formate in 2% formic acid. This is followed by a methanol wash and finally the samples were eluted with 300uL of 5% ammonium hydroxide in methanol. The eluates were then dried to completeness using a SpeedVac (LabConco).

#### High-pH Off-line Fractionation

Dried samples were re-suspended in high pH loading buffer (0.07% vol/vol NH4OH, 0.045% vol/vol FA, 2% vol/vol ACN) and loaded onto a Water’s BEH 1.7 um 2.1mm by 150mm. A Thermo Vanquish was used to carry out the fractionation. Solvent A consisted of 0.0175% (vol/vol) NH4OH, 0.01125% (vol/vol) FA, and 2% (vol/vol) ACN; solvent B consisted of 0.0175% (vol/vol) NH4OH, 0.01125% (vol/vol) FA, and 90% (vol/vol) ACN. The sample elution was performed over a 25 min gradient with a flow rate of 0.6 mL/min. A total of 192 individual equal volume fractions were collected across the gradient and subsequently pooled by concatenation into 96 fractions and dried to completeness using a SpeedVac.

#### Liquid Chromatography Tandem Mass Spectrometry

All fractions were resuspended in an equal volume of loading buffer (0.1% FA, 0.03% TFA, 1% ACN) and analyzed by liquid chromatography coupled to tandem mass spectrometry. Peptide eluents were separated on Water’s CSH column (1.7um resin 150um by 15 cm) by a Vanquish Neo (ThermoFisher Scientific). Buffer A was water with 0.1% (vol/vol) formic acid, and buffer B was 99.9% (vol/vol) acetonitrile in water with 0.1% (vol/vol) formic acid. The gradient was from 3% to 35% solvent B over 17 mins followed by column wash and equilibration for a total of 23 mins. Peptides were monitored on a Orbitrap Astral spectrometer (ThermoFisher Scientific) fitted with a high-field asymmetric waveform ion mobility spectrometry (FAIMS Pro) ion mobility source (ThermoFisher Scientific). Two compensation voltages (CV) of −45 and −60 were chosen for the FAIMS. Each cycle consisted of one full scan acquisition (MS1) with an m/z range of 400-1500 at 120,000 resolution and standard settings and as many tandem (MS/MS) scans in 1.5 seconds. The Astral higher energy collision-induced dissociation (HCD) tandem scans were collected at 35% collision energy with an isolation of 0.5 m/z, an AGC setting of 100%, and a maximum injection time of 20ms. Dynamic exclusion was set to exclude previously sequenced peaks for 30 seconds within a 5-ppm tolerance window.

#### Database search

6958 raw files (across 65 TMT-18 plexes; deposited at www.synapse.org with the SynID: syn59804727) were searched using FragPipe (version 22.0), as described above. The FragPipe pipeline relies on MSFragger (version 4.0)^72,73^ Percolator^74^for peptide identification, MSBooster^75^ and Percolator^74^ for false discovery rate (FDR) filtering and downstream processing. The search was performed with a database of canonical Human proteins downloaded from Uniprot (20,402; accessed 02/11/2019), as well as sequences for specific peptides for APOE4 and 2 alleles^23^ alongside ABETA40 and 42 peptides as described^23^ (total of 20,405 sequences). The workflow used in FragPipe followed default TMT-16 plex parameters, used for both TMT-16 and TMT-18 experimental design. Briefly, precursor mass tolerance was −20 to 20 ppm, fragment mass tolerance of 20 ppm, mass calibration and parameter optimization were selected, and isotope error was set to −1/0/1/2/3. Enzyme specificity was set to semi-tryptic and up to two missing trypsin cleavages were allowed. Peptide length was allowed to range from 7 to 42 and peptide mass from either 200 to 5,000 Da. Variable modifications that were allowed in our search included: oxidation on methionine, N-terminal acetylation on protein, N-terminal acetylation on peptide along with off-target TMT tag modification on Serine, Threonine and Histidine, with a maximum of 3 variable modifications per peptide. Peptide Spectral Matches were validated using Percolator^74^. The FDR threshold was set to 1% and protein and peptide abundances were quantified using Philosopher for downstream analysis. Before performing any abundance analysis, the data from all batches were merged and protein levels were first scaled by dividing each protein intensity by intensity sum of all proteins in each sample followed by multiplying by the maximum protein intensity sum across all nine samples. Instances where the intensity was ‘0’ were treated as ‘missing values’. The searched data matrix has been deposited with ADNI Biofluid Biomarker Core.

#### Tunable Approach for Median Polish of Ratio (TAMPOR)

We used TAMPOR (Tunable Approach for Median Polish of Ratio) to remove technical batch variance in the proteomic data, as previously described^76^. TAMPOR removes inter-batch variance while preserving variance caused by biological changes in the protein abundance values, normalizing to the median of selected samples.

#### Regression of unwanted covariates

The protein abundance matrices are then subjected to non-parametric bootstrap regression by subtracting the trait of interest (post-mortem interval (PMI) of the sample, batch of the sample) multiplied by the median estimated coefficient of fitting for each protein in the log2(abundance) matrix.

#### Differential Protein Expression Analysis

Linear models were used to identify differentially abundant proteins (DAPs) associated with neuroimaging vascular abnormalities, amyloid-β (Aβ) pathology, and cognitive performance. The first set of comparisons focused on DAPs related to vascular abnormalities, including WMH, microbleeds, and infarcts. Microbleeds and infarcts were modeled as binary variables (presence vs. absence), whereas WMH burden was treated as a continuous outcome. Next, differential expression analysis was performed using the Alzheimer’s Disease Assessment Scale–Cognitive Subscale(-Cog11)^93^, a widely used measure of global cognitive function. Separate models contrasting participants negative for amyloid-β pathology (as determined by AV-45 PET scans) with those positive for amyloid-β pathology were performed to investigate proteins specifically associated with amyloid-β. Amyloid-β pathology status was based on the AMYLOID-STATUS variable provided in the ADNI database. Subsequently, differential expression analysis was conducted by dividing participants into clinically diagnosed groups (controls, MCI, and dementia) and performing pairwise comparisons. All results are presented as volcano plots or heatmaps. To account for multiple hypothesis testing, we applied the Benjamini-Hochberg method to adjust p values using the false discovery rate (FDR). Given our goal to broadly characterize protein-level alterations, we considered results significant at P < 0.05.

#### Machine Learning Modelling

We assessed the predictive capability of CSF proteins for neurovascular abnormalities, including WMH, infarcts, and microbleeds. Tailored machine learning pipelines were developed for each outcome, taking into account the distribution characteristics of the target variable and ensuring robust feature selection and classification. For WMH prediction, participants were stratified into quartiles based on total WMH volume. To focus on clinically meaningful extremes, only participants in the top (highest WMH burden) and bottom quartiles (lowest WMH burden) were retained for binary classification (n=384; top quartile: n=192; bottom quartile: n=192). Participants in the middle two quartiles were excluded to reduce ambiguity and improve class separability. This approach allowed us to model clear phenotypic differences in WMH burden. Both infarcts and microbleeds presented class imbalance, with a relatively smaller proportion of participants exhibiting these lesions (microbleeds: n=723, n=194 with microbleeds, n=529 without microbleeds; infarcts: n=1050, n=81 with infarcts, n=969 without infarcts). To address this imbalance, we applied the Synthetic Minority Oversampling Technique (SMOTE)^94^ to the training data only. SMOTE generates synthetic examples of the minority class by interpolating between existing instances, thus enhancing the classifier’s ability to detect rare events without overfitting to duplicated data. During model development for each of the three outcomes, we applied support vector machine-based forward feature selection (SVM-FFS)^95^ to identify informative peptides, rather than using all available features. Beginning with no features, the algorithm iteratively added the features that most improved classification performance, as determined by cross-validation accuracy on the training set. This wrapper-based approach enabled us to identify a compact, high-performing feature set from a potentially high-dimensional dataset. Following feature selection, SVM classifier was trained using the selected features. SVMs are well-suited for high-dimensional biomedical data and provide strong performance in capturing complex, potentially non-linear relationships, especially when appropriate kernels are used. Separate SVM classifiers were trained for WMH, infarcts, and microbleeds tasks. The dataset was split into 75% training and 25% test sets. All feature selection and hyperparameter tuning (e.g., kernel type, regularization parameter C, and kernel coefficient gamma) were conducted exclusively within the training data using cross-validation and grid search. The independent test set was held out entirely during model development and used solely to evaluate final performance. Machine learning analyses were conducted in Python (version 3.9), using libraries such as scikit-learn for model development and evaluation, and imbalanced-learn for SMOTE implementation. Only proteins with data available for more than 95% of participants were included in this analysis. Remaining missing values were imputed using the mean value of the corresponding clinical group (e.g., control, MCI, or dementia). Agreement between observed versus predicted class labels was assessed using area-under-the-receiver-operating-characteristic (ROC) curve. Optimal thresholds along the ROC were defined using the Youden index^96^. ROC curve analyses were performed in R using the pROC package to assess model performance and compute the area under the curve (AUC). Feature selection, model training, and validation on an independent test set were performed separately for each of the three outcomes.

#### Cerebrovascular module association to vascular imaging abnormalities

To evaluate the relationship between cerebrovascular network modules and MRI-based vascular abnormalities, we analyzed CSF proteins correlated to WMH, microbleeds, infarcts, and AV45. Gene products and their associated p values were tested for enrichment within cerebrovascular network modules using a permutation-based framework (10,000 permutations) implemented in R, with exact p-values derived using the ‘permp’ function from the statmod package. Module-specific enrichment was quantified by calculating Z scores, as previously described^12^ using the enrichment analysis applied to GWAS-derived gene-level statistics (available at https://www.github.com/edammer/MAGMA.SPA).

#### Bio-Layer Interferometry

To determine whether CRIP1 binds to Aβ 1-40 and Aβ 1-42, we performed Bio-Layer Interferometry (BLI) using BLITZ instrument (BLItz™ Protein Detection System). Initially, the baseline reading was measured with OCTET kinetics buffer. After baseline reading, we loaded 10 uM Biotinylated Aβ40 (A-111-2) and Aβ42 (A1117-1) (10 μM) from rPeptide on an Octet® High Precision Streptavidin (SAX) Biosensors (18-5117), respectively for 120 s. Next, the loaded biosensors were suspended (30 s) into OCTET kinetics buffer to remove any unbound Aβ40 and Aβ42. Further, the CRIP1 at various concentrations (0.25 mM, 0.5 mM, 1 mM, 9.1 mM) were associated (120 s). The dissociation was further performed for 120 s in OCTET kinetics buffer to remove unbound CRIP1. The background binding signal were subtracted from all the experiments. The analysis of global binding kinetics was performed using BLITZ software.

#### Thioflavine-T aggregation of Aβ with CRIP1

Recombinant human Aβ1-40 (10 µM, from rPeptide A-1180-) or Aβ 1-42 (10 µM) (rPeptide A-1170-1) was incubated in 50 mM Na Phosphate Buffer (pH 7.4), 5 mM TCEP and 40 µM ThT or in Tris-buffered Saline (TBS; 150 mM NaCl, 50 mM Tris-HCl, pH 7.6), 5 mM TCEP and 20 µM ThT, respectively in the presence or absence of purified recombinant human CRIP1 His protein (1, 10 and 50 nM) from Novus Biologicals (NBP2-22979). The final volume within each well was 100 µL. The assay was conducted in quadruplicates using chilled (4°C) 96 well black clear bottom plates (Corning, #3904). Fluorescence was captured at 420 Ex, 480 Em for 20 hours at 15 min intervals at 37°C using Synergy H1 (Biotek) microplate reader. ThT alone (20 µM) was measured and subtracted as background fluorescence. Fluorescence intensities were graphed using GraphPad Prism.

#### CSF pQTL mapping

As previously described^44^, pQTL mapping was conducted using SNPs with a minor allele frequency (MAF) ≥ 0.05, defining cis pQTLs as variants within 1 Mb of the corresponding protein-coding gene and trans pQTLs as variants outside this window. Linear models were fitted in PLINK^97^ with SNP genotype as the predictor and normalized protein expression as the outcome, adjusting for sex, 10 genetic principal components, and genetic data source (WGS vs. array). Significance thresholds were p < 5×10^-8^ for cis pQTLs. To account for linkage disequilibrium among significant pQTLs, PLINK’s ‘clump’ function was used to identify independent index variants with pairwise r^2^ < 0.1 within a 5 Mb window. The genomic inflation factor (λ) was estimated for each analysis, with a median λ of 1.002 (range 0.975–1.028).

#### Integrated analysis of CSF proteomics with AD GWAS data

The integration of CSF proteomic and genetic data with AD GWAS^43^ resulted from using two complementary approaches. A proteome-wide association study (PWAS) was performed with the FUSION pipeline^98^, restricting analyses to SNPs from the European LD reference panel, estimating SNP-based heritability for each protein, and designating those with p < 0.01 as heritable. For heritable proteins, predictive models (BLUP, LASSO, elastic net, BSLMM) were trained using SNPs within ±500 kb of the gene, SNP weights from the best-performing model were retained, and these were integrated with AD GWAS summary statistics to identify cis-regulated proteins associated with AD (FDR < 0.05). Summary data–based Mendelian Randomization (SMR)^99^ was applied to PWAS-significant proteins, combining CSF pQTL and AD GWAS data to test whether protein abundance mediated SNP–trait associations, and HEIDI^99^ testing was used to exclude associations likely due to linkage disequilibrium (HEIDI p < 0.05). Proteins were considered causally implicated if they satisfied all criteria: PWAS FDR < 0.05, SMR p < 0.05, HEIDI p > 0.05, and consistent effect directions between PWAS and SMR. All genomic coordinates were mapped to GRCh38.

## Author Contributions

Conceptualization: AMW and NTS; Formal analysis: A.M.W., S.R., E.B.D., L.Y., A.S., Y.L., S.O., A.P.W., T.S.W., and N.T.S; Funding Acquisition: A.M.W., S.R., G.R.H., A.I.L., and N.T.S.; Investigation: A.M.W., F.S., J.G., D.M.D., F.W., K.M.K., Z.T.M., N.T.S.; Methodology: A.M.W., S.R., E.B.D., L.Y., A.N.T., A.P.W., T.S.W., and N.T.S.; Project administration: A.M.W., E.J.F., N.T.S; Resources: M.G., S.R., J.A.S., E.B.L., D.A.B., J.J.L., T.D., E.C.B.J., V.M., Z.T.M., and N.T.S.; Software: S.R., E.B.D., L.Y., A.P.W., T.S.W., and N.T.S.; Supervision: A.I.L., N.T.S; Visualization: A.M.W., S.R., and N.T.S.;Writing-original draft: A.M.W., N.T.S.; Writing-review & editing: A.M.W., S.R., E.B.D., and N.T.S; All authors read and approved the final manuscript.

## Disclosures

N.T.S, A.I.L and D.M.D. are co-founders and consultants of Emtherapro. D.M.D. and N.T.S are co-founders of Arc Proteomics. N.T.S, and Z.T.M are co-founders of StitchRx. N.T.S has consulted for AbbVie, Eisai, Trace Neuroscience, and Arrowhead Pharmaceuticals.

## Data Availability

Raw mass spectrometry data and pre- and post-processed (brain and CSF) protein abundance data and case traits related to this manuscript are available at https://www.synapse.org/Synapse:syn69962262. The AMP-AD Knowledge Portal is a platform for accessing data, analyses and tools generated by the AMP-AD Target Discovery Program and other programs supported by the National Institute on Aging to enable open-science practices and accelerate translational learning. The data, analyses and tools are shared early in the research cycle without a publication embargo on secondary use. Data are available for general research use according to the following requirements for data access and data attribution in the ADNI Biofluid Biomarker Core.

## Acknowledgements Section

Brain tissues were provided by the Emory University and University of Pennsylvania School of Medicine Brain Banks. This study was supported by the following National Institutes of Health (NIH) funding mechanisms: U01AG061357 (A.I.L. and N.T.S.), RF1AG062181 (N.T.S.), F32AG081135 (A.M.W.), P30AG066511 (A.I.L.), RF1NS139948 (N.T.S. and G.H.), R01AG074569 (T.G.), and R01AG075820 (N.T.S. and S.R.); as well as the Foundation for the National Institutes of Health AMP-AD 2.0 grant and a grant from the Alzheimer’s Association (ABA-22-974673).

Data collection and sharing for this project was funded by the Alzheimer’s Disease Neuroimaging Initiative (ADNI) (National Institutes of Health Grant U01 AG024904) and DOD ADNI (Department of Defense award number W81XWH-12-2-0012). ADNI is funded by the National Institute on Aging, the National Institute of Biomedical Imaging and Bioengineering, and through generous contributions from the following: AbbVie, Alzheimer’s Association; Alzheimer’s Drug Discovery Foundation; Araclon Biotech; BioClinica, Inc.; Biogen; Bristol-Myers Squibb Company; CereSpir, Inc.; Cogstate; Eisai Inc.; Elan Pharmaceuticals, Inc.; Eli Lilly and Company; EuroImmun; F. Hoffmann-La Roche Ltd and its affiliated company Genentech, Inc.; Fujirebio; GE Healthcare; IXICO Ltd.; Janssen Alzheimer Immunotherapy Research & Development, LLC.; Johnson & Johnson Pharmaceutical Research & Development LLC.; Lumosity; Lundbeck; Merck & Co., Inc.; Meso Scale Diagnostics, LLC.; NeuroRx Research; Neurotrack Technologies; Novartis Pharmaceuticals Corporation; Pfizer Inc.; Piramal Imaging; Servier; Takeda Pharmaceutical Company; and Transition Therapeutics. The Canadian Institutes of Health Research is providing funds to support ADNI clinical sites in Canada. Private sector contributions are facilitated by the Foundation for the National Institutes of Health (www.fnih.org). The grantee organization is the Northern California Institute for Research and Education, and the study is coordinated by the Alzheimer’s Therapeutic Research Institute at the University of Southern California. ADNI data are disseminated by the Laboratory for Neuro Imaging at the University of Southern California.

---

**Figure S1.**
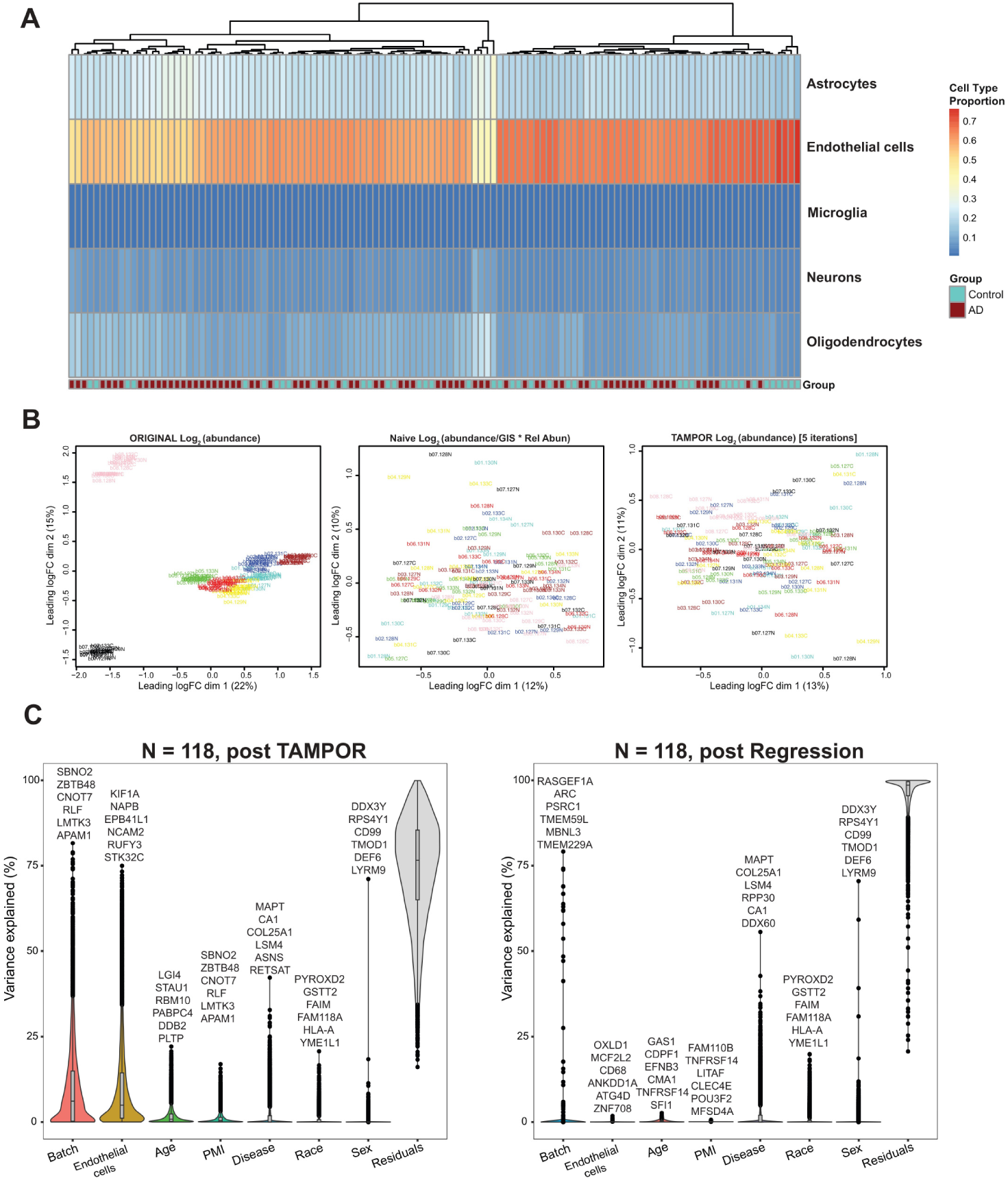
Assessment of cell type enrichment, batch effects, and co-variates on the cerebrovascular proteome. (A) Heatmap illustrating core brain cell type contribution to cerebrovascular proteome. Proportions of the five cell types are defined via reprocessing of Darmanis, Nowakowski, and Zhong single-cell datasets as curated with the EnsDeconv R package, which calculated cell type proportions by ensemble deconvolution of cerebrovascular proteome. Dark red and orange indicate high enrichment of endothelial proteins to proportions above 70 percent, and dark blue indicates the lowest enrichment of cell-specific proteins in the preparations. (B) Multidimensional scaling (MDS) illustrating TMT-MS batch correction. Log_2_ abundance and log_2_ abundance divided by the global internal standard (GIS) are shown. (C) Variance partition plots were used to visualize the percent variance of each protein in the dataset co-varying with batch, endothelial cell type proportions, age, postmortem interval (PMI), disease, race, and sex. The matrix was subjected to bootstrap regression (right) to remove variance due to age, PMI, and endothelial cell type proportions.

**Figure S2.**
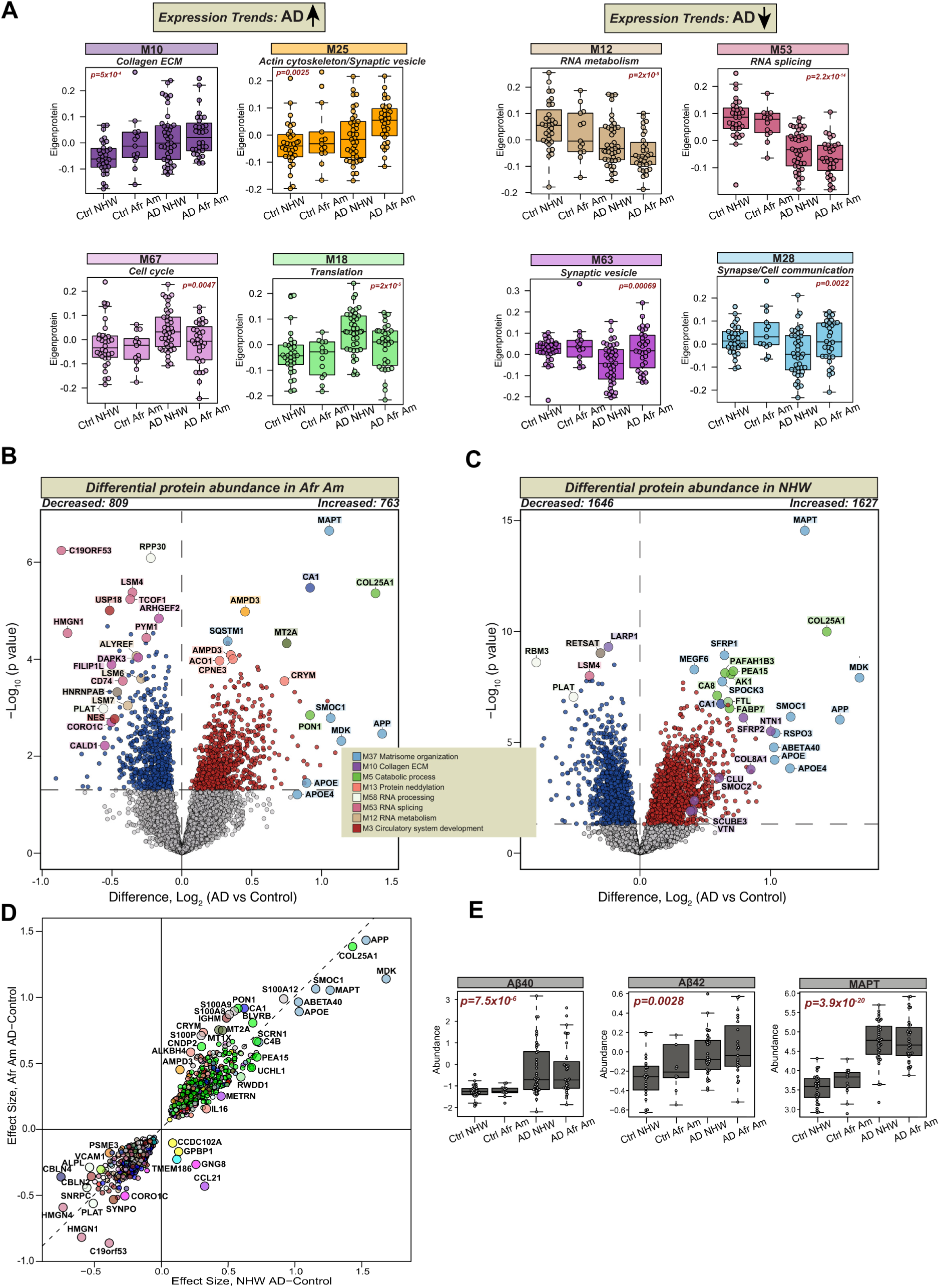
Race-specific cerebrovascular proteomic signatures in Alzheimer’s Disease reveal distinct and shared pathways in African American and Non-Hispanic White individuals. (A) Frontal cortical brain samples were collected from AD and control individuals (Emory: N= 11 African American (Afr Am), N= 9 Non-Hispanic Whites (NHW); UPenn: N=2 Afr Am, N=23 NHW) or Alzheimer’s disease (Emory: N= 32 Afr Am, N= 29 NHW; UPenn: N=12). Module eigenprotein (ME) levels by race were presented for protein modules showing a significant change between groups. MEs were obtained by using one-way ANOVA test and p values are provided for each module. Box plots represent median, 25^th^ and 75^th^ percentile while box whiskers encompass actual data points up to 1.5 times the nearest interquartile range. Gene ontology analysis was used to assign a biological process to each module. (B-C) Volcano plots showing differential abundance of proteins measured in cerebrovasculature of African American (Afr Am) (b) or non-Hispanic White (NHW) (c), individuals between control and AD groups. The x axis illustrates the log_2_ fold change (AD vs. Control), while the y axis represents the −log_10_ statistical p value calculated for all proteins in each pairwise group, obtained as Tukey post-hoc test p values following one-way analysis of variance (ANOVA), except for imprecisely calculated Tukey values below 10^−8.5^ which underwent more precise and stringent Bonferroni post-hoc correction of a two-tailed unequal variance t-test. Proteins significantly elevated in Afr Am with AD (N=763) (b) or NHW (N=1627) (c) are shown in red whereas those significantly decreased in Afr Am AD (N = 809) (b) or NHW AD (N = 1646) (c) are depicted in blue (p < 0.05). Proteins of interest are shown as enlarged dots and shaded according to the color of their module membership. (D) Scatterplot displaying a Pearson correlation between log_2_ effect size (AD vs. Control) of significantly altered proteins in cerebrovasculature of NHW and Afr Am cases. The significance of Pearson correlation was determined by Student’s t-test for significance of correlation implemented in the WGCNA R package. (E) TMT-MS quantified levels of amyloid-β (Aβ)40, Aβ42, and microtubule associated protein tau (MAPT) in cerebrovascular preparations across ethnoracial and diagnosis groups. Box plots represent median, 25th and 75th percentiles while box whiskers extend to non-outlier measurements up to 1.5 times each nearest interquartile range.

**Figure S3.**
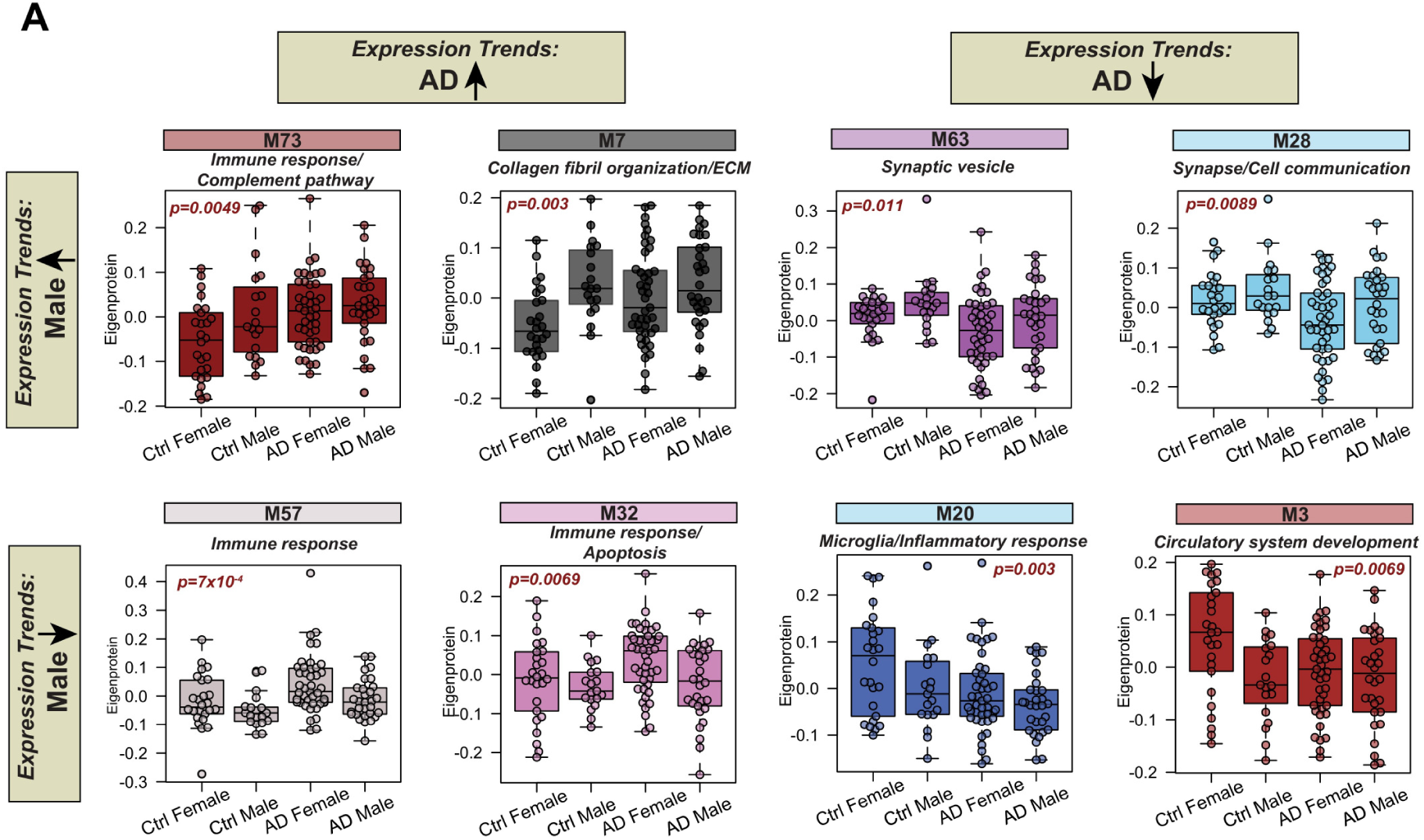
Sex- and diagnosis-associated variation in module abundance. (A-B) Boxplots of selected modules display module eigenprotein level by sex (A) as well as sex and diagnosis (B). Box plots represent median, 25^th^ and 75^th^ percentile while box whiskers encompass actual data points up to 1.5 times the nearest interquartile range. Principal biology of each module was assessed by gene ontology analysis.

**Figure S4.**
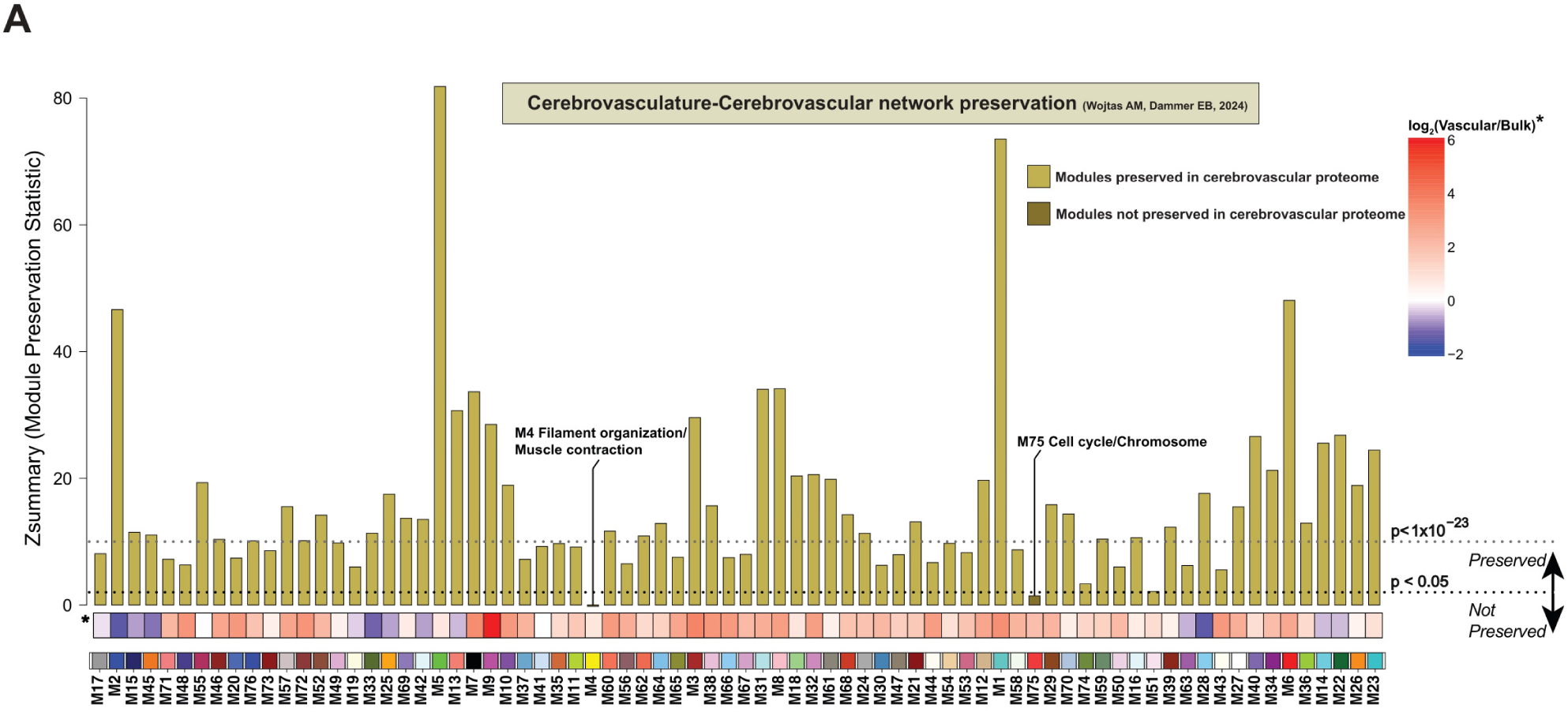
Modules associated with vascular biology are preserved in independent cerebrovascular proteome. (A) Module preservation of the current TMT cerebrovascular network into the previously published cerebrovascular network (*Wojtas AM, Dammer EB, 2024*). Z_summary_ score greater than or equal to 1.96 (or p< 0.05) indicated module preservation, whereas Z_summary_ score less than 1.96 showed lack of preservation. Bar colors indicate the status of preservation.

**Figure S5.**
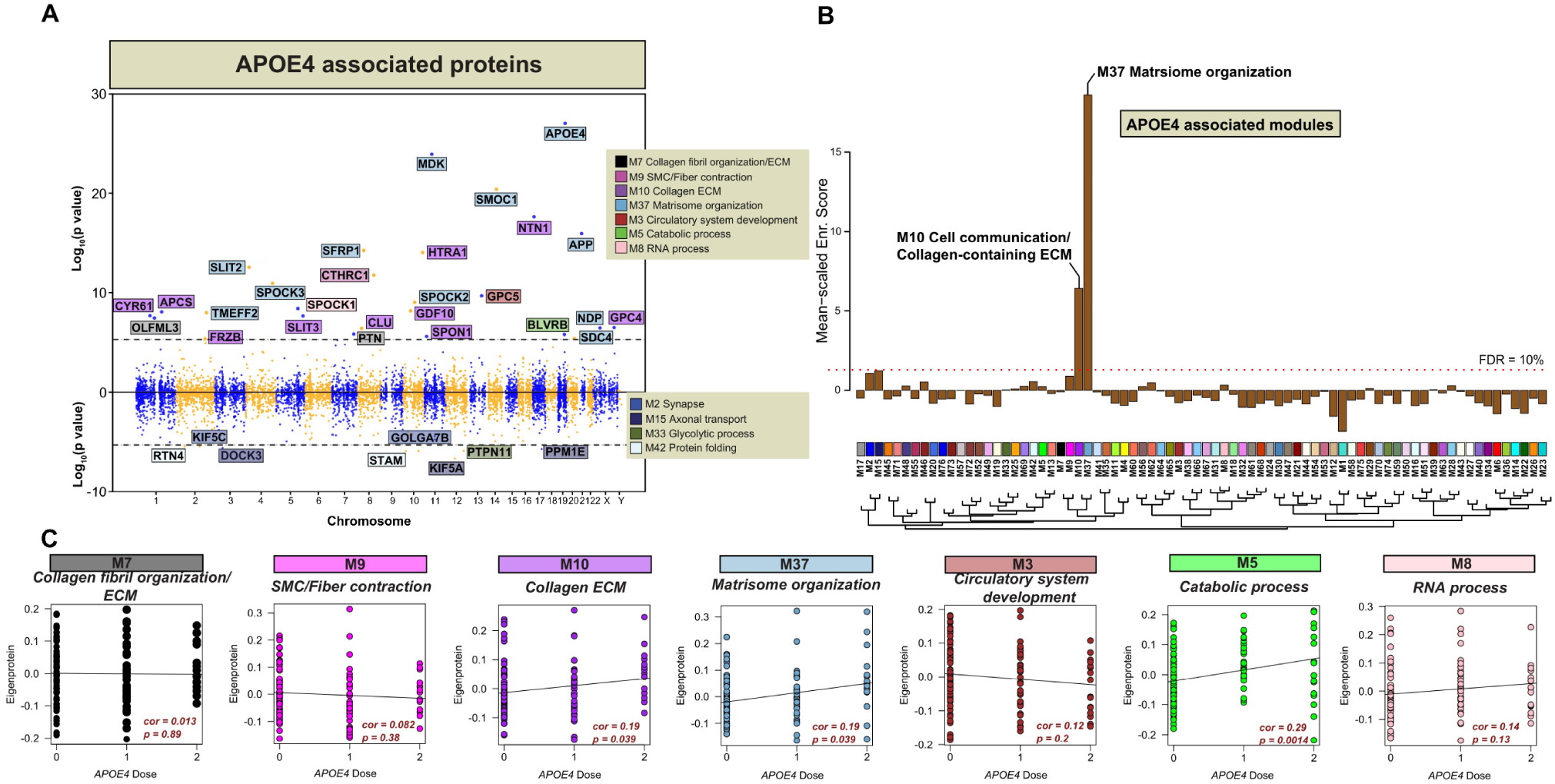
*APOE* genotype is associated with extracellular matrix proteins. (A) 10,030 proteins measured in bulk tissue from ROSMAP cases (N=873). X-axis shows chromosome location for individual proteins and y-axis depicts positive and negative log_10_ *p values*. Proteins that survived Bonferroni correction are labeled. The colors indicate module membership in cerebrovascular network. (B) Cerebrovascular module association to APOE4. (C) Correlation plots of selected modules to *APOE4* dose (*APOE2/2,* N=1*; APOE2/3,* N=4*; APOE3/3,* N=44*; APOE2/4,* N=2*; APOE3/4,* N=36*; APOE4/4,* N=16). Correlation and p value are shown for each module.

**Figure S6.**
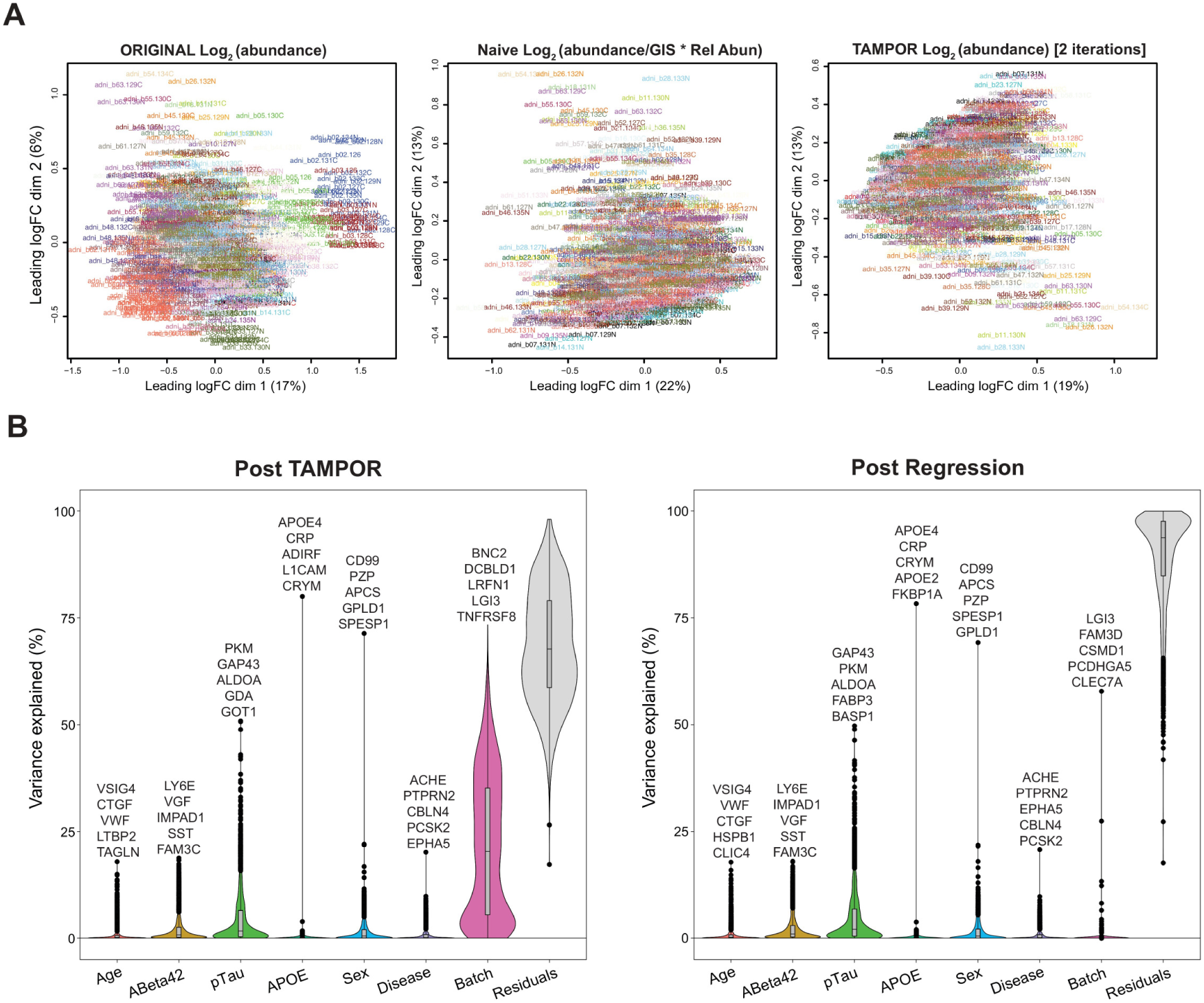
Quality control of the ADNI CSF proteome. (A) Multidimensional scaling (MDS) illustrating TMT-MS batch correction. Log_2_ abundance, log_2_ abundance divided by the global internal standard (GIS), and TAMPOR are shown. (B) Variance partition plots were used to visualize the percent variance of each protein in the dataset co-varying with batch, age and sex. The matrix was subjected to bootstrap regression (right) to remove variance due to batch.

**Figure S7.**
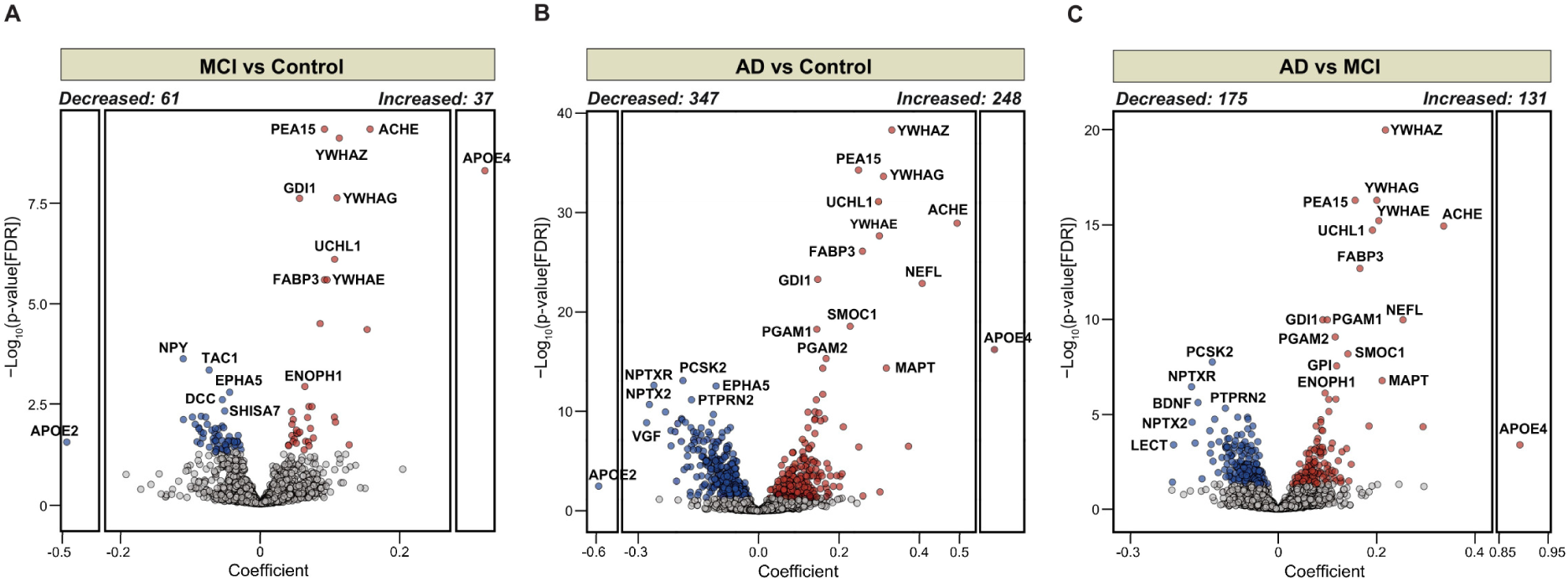
Differential abundance of CSF proteins in ADNI across clinical stages of dementia. (A-C) Volcano plots showing differential abundance of proteins at P_FDR_<0.05 between clinically diagnosed control and MCI (Increased: 37, Decreased: 61) A), control and AD (Increased: 248, Decreased: 347) (B), and MCI and AD (Increased: 131, Decreased: 175) (C) (Controls: N=377, MCI: N=563, AD: N=164). The x axis represents the mean difference while the y axis shows the -log_10_ statistical *p* value calculated for all proteins in each group. *P* values were obtained from Student’s t test. Proteins significantly increased are shown in red, whereas proteins significantly decreased are highlighted in blue. Proteins with unchanged levels are represented in grey.

**Figure S8.**
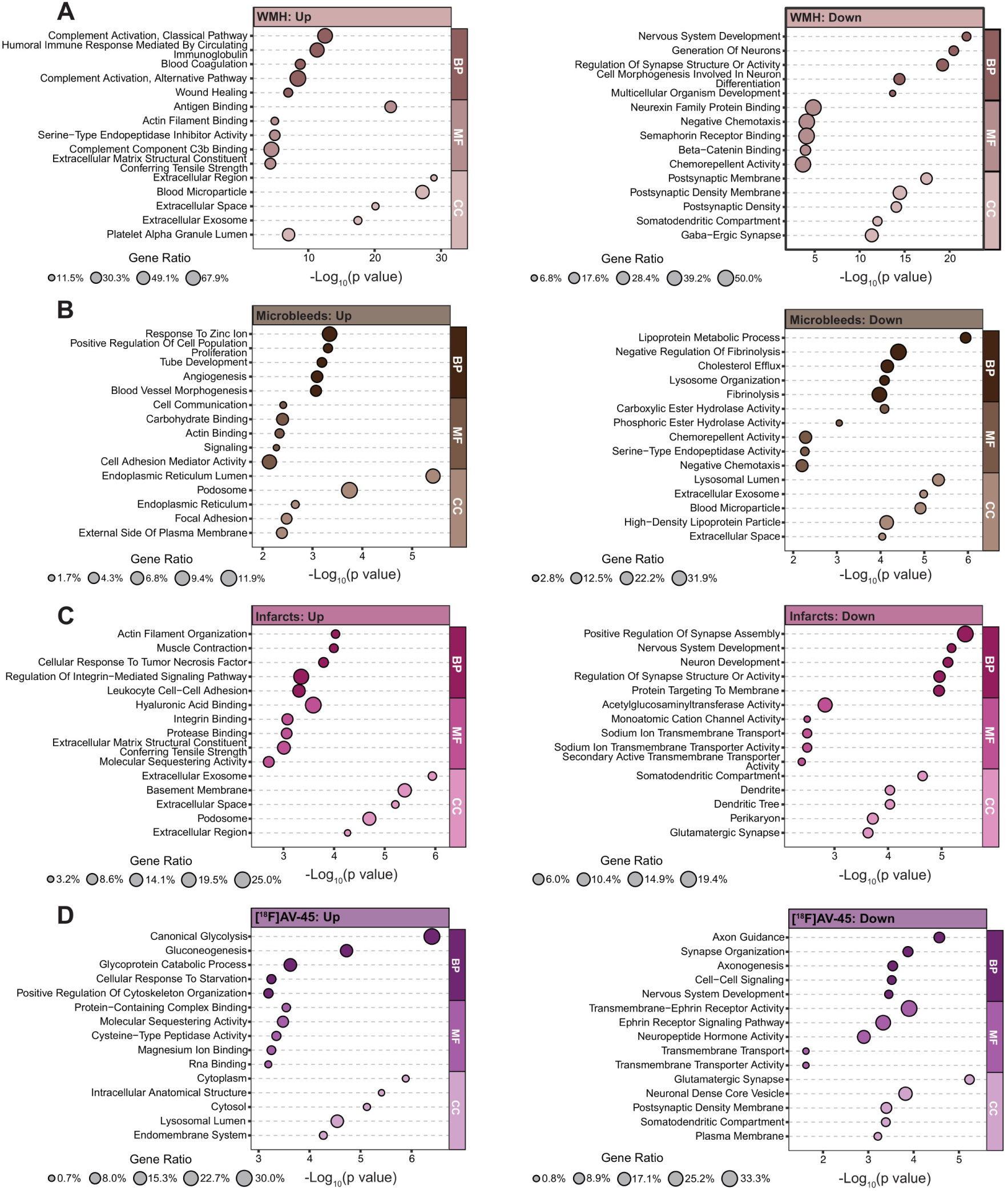
Biological processes underlying neuroimaging-based vascular abnormalities. (A) Gene ontology analysis was performed to identify biological processes associated with each neurovascular manifestations (WMH, microbleeds, infarcts) as well as AV-45 using the proteins significantly associated with each imaging phenotype, as shown in Fig. 4. Upregulated and downregulated pathways are separately shown to highlight the directionality of the associations. Sample sizes were: WMH (N = 775), infarcts (N = 1,050), microbleeds (N = 732), and AV-45 (N = 745).

**Figure S9.**
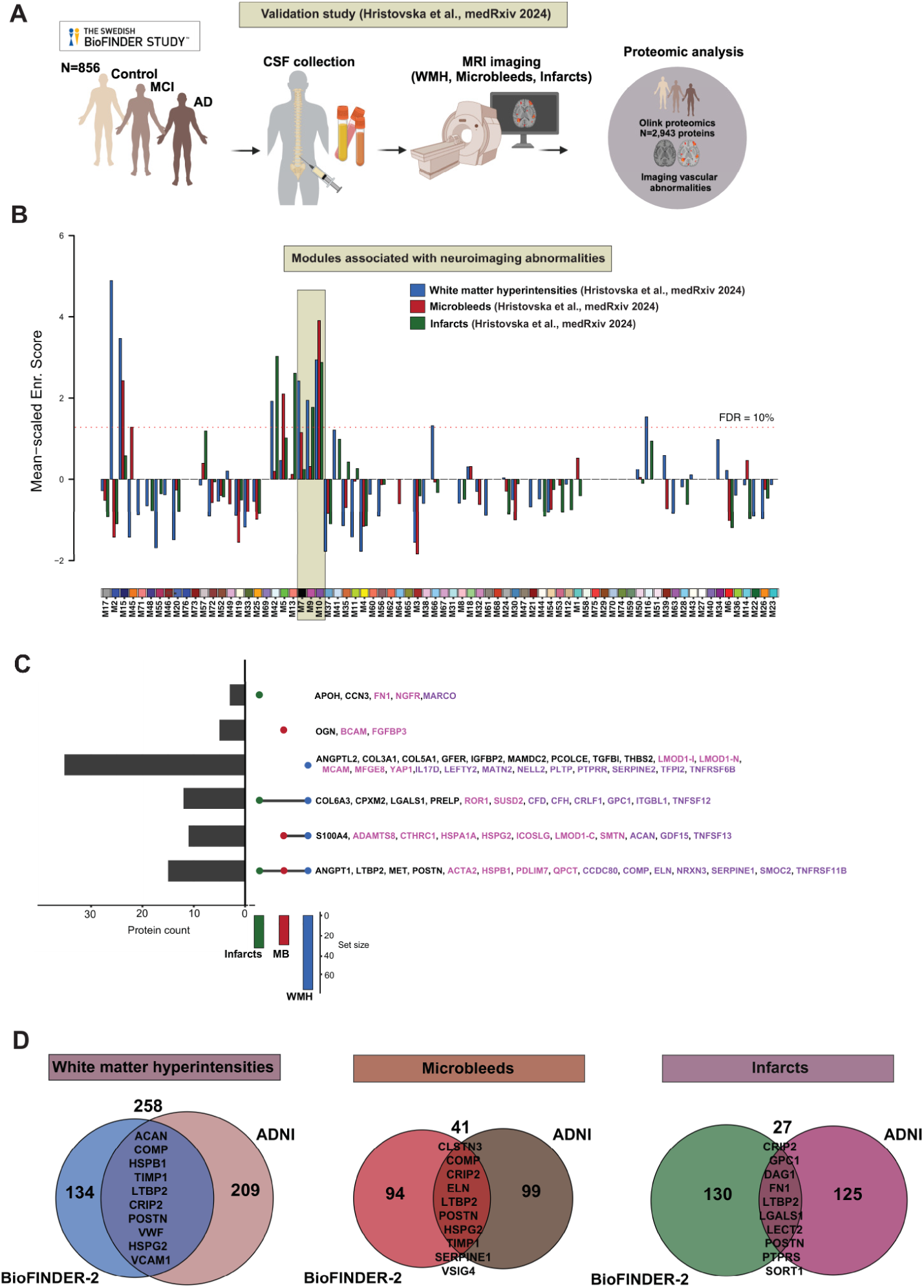
Cross-cohort validation of cerebrovascular protein modules linked to imaging markers and CAA severity in BioFINDER-2 and ADNI. (A) Schematic representation of workflow in the Swedish BioFINDER study. (B) Cerebrovascular network modules enriched for CSF proteins associated with imaging abnormalities from BioFINDER-2 cohort. (C) UpSet plot illustrating proteins associated with WMH, microbleeds, and infarcts, highlighting the shared molecular signature across fluid-based markers of neurovascular injury in BioFINDER-2 cohort. (D) Venn diagrams showing the overlap between BioFINDER-2 and ADNI cohorts in the number of proteins associated with WMH, microbleeds, and infarcts. Top 10 overlapping proteins between datasets are shown in the boxes. An overlapping set of 886 proteins between ADNI and BioFINDER-2 was used in panels c and d, with significance defined as P<0.05 in both the ADNI and BioFINDER-2 cohorts.

## Notes

### Author Declarations

ADNI has been approved by an Institutional Review Board. Religious Orders Study (ROS) or the Rush Memory and Aging Project (MAP), two ongoing longitudinal cohort studies of aging and dementia approved by an Institutional Review Board of Rush University Medical Center. Our study was approved by local IRB review committees at Emory and UPenn.

### Summary of Updates

We have now included the figures embedded in the body of the manuscript and made minor edits to the results section.

## References

1. Inoue, Y., Shue, F., Bu, G., and Kanekiyo, T. (2023). Pathophysiology and probable etiology of cerebral small vessel disease in vascular dementia and Alzheimer’s disease. Mol Neurodegener 18, 46. 10.1186/s13024-023-00640-5.

2. Gladman, J.T., Corriveau, R.A., Debette, S., Dichgans, M., Greenberg, S.M., Sachdev, P.S., Wardlaw, J.M., and Biessels, G.J. (2019). Vascular contributions to cognitive impairment and dementia: Research consortia that focus on etiology and treatable targets to lessen the burden of dementia worldwide. Alzheimers Dement (N Y) 5, 789–796. 10.1016/j.trci.2019.09.017.

3. Morrison, C., Dadar, M., Villeneuve, S., and Collins, D.L. (2022). White matter lesions may be an early marker for age-related cognitive decline. Neuroimage Clin 35, 103096. 10.1016/j.nicl.2022.103096.

4. Greenberg, S.M., Bacskai, B.J., Hernandez-Guillamon, M., Pruzin, J., Sperling, R., and van Veluw, S.J. (2020). Cerebral amyloid angiopathy and Alzheimer disease - one peptide, two pathways. Nat Rev Neurol 16, 30–42. 10.1038/s41582-019-0281-2.

5. Jakel, L., De Kort, A.M., Klijn, C.J.M., Schreuder, F., and Verbeek, M.M. (2022). Prevalence of cerebral amyloid angiopathy: A systematic review and meta-analysis. Alzheimers Dement 18, 10–28. 10.1002/alz.12366.

6. Sweeney, M.D., Kisler, K., Montagne, A., Toga, A.W., and Zlokovic, B.V. (2018). The role of brain vasculature in neurodegenerative disorders. Nat Neurosci 21, 1318–1331. 10.1038/s41593-018-0234-x.

7. Reijmer, Y.D., van Veluw, S.J., and Greenberg, S.M. (2016). Ischemic brain injury in cerebral amyloid angiopathy. J Cereb Blood Flow Metab 36, 40–54. 10.1038/jcbfm.2015.88.

8. Zhang, Y., Chen, H., Li, R., Sterling, K., and Song, W. (2023). Amyloid beta-based therapy for Alzheimer’s disease: challenges, successes and future. Signal Transduct Target Ther 8, 248. 10.1038/s41392-023-01484-7.

9. Sevigny, J., Chiao, P., Bussiere, T., Weinreb, P.H., Williams, L., Maier, M., Dunstan, R., Salloway, S., Chen, T., Ling, Y., et al. (2016). The antibody aducanumab reduces Abeta plaques in Alzheimer’s disease. Nature 537, 50–56. 10.1038/nature19323.

10. Lacorte, E., Ancidoni, A., Zaccaria, V., Remoli, G., Tariciotti, L., Bellomo, G., Sciancalepore, F., Corbo, M., Lombardo, F.L., Bacigalupo, I., et al. (2022). Safety and Efficacy of Monoclonal Antibodies for Alzheimer’s Disease: A Systematic Review and Meta-Analysis of Published and Unpublished Clinical Trials. J Alzheimers Dis 87, 101–129. 10.3233/JAD-220046.

11. Johnson, E.C.B., Dammer, E.B., Duong, D.M., Ping, L., Zhou, M., Yin, L., Higginbotham, L.A., Guajardo, A., White, B., Troncoso, J.C., et al. (2020). Large-scale proteomic analysis of Alzheimer’s disease brain and cerebrospinal fluid reveals early changes in energy metabolism associated with microglia and astrocyte activation. Nat Med 26, 769–780. 10.1038/s41591-020-0815-6.

12. Johnson, E.C.B., Carter, E.K., Dammer, E.B., Duong, D.M., Gerasimov, E.S., Liu, Y., Liu, J., Betarbet, R., Ping, L., Yin, L., et al. (2022). Large-scale deep multi-layer analysis of Alzheimer’s disease brain reveals strong proteomic disease-related changes not observed at the RNA level. Nat Neurosci 25, 213–225. 10.1038/s41593-021-00999-y.

13. Higginbotham, L., Ping, L., Dammer, E.B., Duong, D.M., Zhou, M., Gearing, M., Hurst, C., Glass, J.D., Factor, S.A., Johnson, E.C.B., et al. (2020). Integrated proteomics reveals brain-based cerebrospinal fluid biomarkers in asymptomatic and symptomatic Alzheimer’s disease. Sci Adv 6. 10.1126/sciadv.aaz9360.

14. Johnson, E.C.B., Bian, S., Haque, R.U., Carter, E.K., Watson, C.M., Gordon, B.A., Ping, L., Duong, D.M., Epstein, M.P., McDade, E., et al. (2023). Cerebrospinal fluid proteomics define the natural history of autosomal dominant Alzheimer’s disease. Nat Med 29, 1979–1988. 10.1038/s41591-023-02476-4.

15. Zellner, A., Muller, S.A., Lindner, B., Beaufort, N., Rozemuller, A.J.M., Arzberger, T., Gassen, N.C., Lichtenthaler, S.F., Kuster, B., Haffner, C., and Dichgans, M. (2022). Proteomic profiling in cerebral amyloid angiopathy reveals an overlap with CADASIL highlighting accumulation of HTRA1 and its substrates. Acta Neuropathol Commun 10, 6. 10.1186/s40478-021-01303-6.

16. Leitner, D., Kavanagh, T., Kanshin, E., Balcomb, K., Pires, G., Thierry, M., Suazo, J.I., Schneider, J., Ueberheide, B., Drummond, E., and Wisniewski, T. (2024). Differences in the cerebral amyloid angiopathy proteome in Alzheimer’s disease and mild cognitive impairment. Acta Neuropathol 148, 9. 10.1007/s00401-024-02767-1.

17. Wojtas, A.M., Dammer, E.B., Guo, Q., Ping, L., Shantaraman, A., Duong, D.M., Yin, L., Fox, E.J., Seifar, F., Lee, E.B., et al. (2024). Proteomic changes in the human cerebrovasculature in Alzheimer’s disease and related tauopathies linked to peripheral biomarkers in plasma and cerebrospinal fluid. Alzheimers Dement 20, 4043–4065. 10.1002/alz.13821.

18. Charidimou, A., Boulouis, G., Frosch, M.P., Baron, J.C., Pasi, M., Albucher, J.F., Banerjee, G., Barbato, C., Bonneville, F., Brandner, S., et al. (2022). The Boston criteria version 2.0 for cerebral amyloid angiopathy: a multicentre, retrospective, MRI-neuropathology diagnostic accuracy study. Lancet Neurol 21, 714–725. 10.1016/S1474-4422(22)00208-3.

19. Knudsen, K.A., Rosand, J., Karluk, D., and Greenberg, S.M. (2001). Clinical diagnosis of cerebral amyloid angiopathy: validation of the Boston criteria. Neurology 56, 537–539. 10.1212/wnl.56.4.537.

20. Greenberg, S.M., and Charidimou, A. (2018). Diagnosis of Cerebral Amyloid Angiopathy: Evolution of the Boston Criteria. Stroke 49, 491–497. 10.1161/STROKEAHA.117.016990.

21. Thal, D.R., Rub, U., Orantes, M., and Braak, H. (2002). Phases of A beta-deposition in the human brain and its relevance for the development of AD. Neurology 58, 1791–1800. 10.1212/wnl.58.12.1791.

22. Yang, A.C., Vest, R.T., Kern, F., Lee, D.P., Agam, M., Maat, C.A., Losada, P.M., Chen, M.B., Schaum, N., Khoury, N., et al. (2022). A human brain vascular atlas reveals diverse mediators of Alzheimer’s risk. Nature 603, 885–892. 10.1038/s41586-021-04369-3.

23. Seifar, F., Fox, E.J., Shantaraman, A., Liu, Y., Dammer, E.B., Modeste, E., Duong, D.M., Yin, L., Trautwig, A.N., Guo, Q., et al. (2024). Large-scale deep proteomic analysis in Alzheimer’s disease brain regions across race and ethnicity. Alzheimers Dement 20, 8878–8897. 10.1002/alz.14360.

24. Kamara, D.M., Gangishetti, U., Gearing, M., Willis-Parker, M., Zhao, L., Hu, W.T., and Walker, L.C. (2018). Cerebral Amyloid Angiopathy: Similarity in African-Americans and Caucasians with Alzheimer’s Disease. J Alzheimers Dis 62, 1815–1826. 10.3233/JAD-170954.

25. Wilkins, C.H., Grant, E.A., Schmitt, S.E., McKeel, D.W., and Morris, J.C. (2006). The neuropathology of Alzheimer disease in African American and white individuals. Arch Neurol 63, 87–90. 10.1001/archneur.63.1.87.

26. Jansen, I.E., Savage, J.E., Watanabe, K., Bryois, J., Williams, D.M., Steinberg, S., Sealock, J., Karlsson, I.K., Hagg, S., Athanasiu, L., et al. (2019). Genome-wide meta-analysis identifies new loci and functional pathways influencing Alzheimer’s disease risk. Nat Genet 51, 404–413. 10.1038/s41588-018-0311-9.

27. Strittmatter, W.J., Saunders, A.M., Schmechel, D., Pericak-Vance, M., Enghild, J., Salvesen, G.S., and Roses, A.D. (1993). Apolipoprotein E: high-avidity binding to beta-amyloid and increased frequency of type 4 allele in late-onset familial Alzheimer disease. Proc Natl Acad Sci U S A 90, 1977–1981. 10.1073/pnas.90.5.1977.

28. Bell, R.D., Winkler, E.A., Singh, I., Sagare, A.P., Deane, R., Wu, Z., Holtzman, D.M., Betsholtz, C., Armulik, A., Sallstrom, J., et al. (2012). Apolipoprotein E controls cerebrovascular integrity via cyclophilin A. Nature 485, 512–516. 10.1038/nature11087.

29. Montagne, A., Nation, D.A., Sagare, A.P., Barisano, G., Sweeney, M.D., Chakhoyan, A., Pachicano, M., Joe, E., Nelson, A.R., D’Orazio, L.M., et al. (2020). APOE4 leads to blood-brain barrier dysfunction predicting cognitive decline. Nature 581, 71–76. 10.1038/s41586-020-2247-3.

30. Hawkes, C.A., Sullivan, P.M., Hands, S., Weller, R.O., Nicoll, J.A., and Carare, R.O. (2012). Disruption of arterial perivascular drainage of amyloid-beta from the brains of mice expressing the human APOE epsilon4 allele. PLoS One 7, e41636. 10.1371/journal.pone.0041636.

31. Fu, Y., Zhao, J., Atagi, Y., Nielsen, H.M., Liu, C.C., Zheng, H., Shinohara, M., Kanekiyo, T., and Bu, G. (2016). Apolipoprotein E lipoprotein particles inhibit amyloid-beta uptake through cell surface heparan sulphate proteoglycan. Mol Neurodegener 11, 37. 10.1186/s13024-016-0099-y.

32. Oveisgharan, S., Yu, L., de Paiva Lopes, K., Tasaki, S., Wang, Y., Menon, V., Schneider, J.A., Seyfried, N.T., and Bennett, D.A. (2024). Proteins linking APOE varepsilon4 with Alzheimer’s disease. Alzheimers Dement 20, 4499–4511. 10.1002/alz.13867.

33. Shen, Y., Timsina, J., Heo, G., Beric, A., Ali, M., Wang, C., Yang, C., Wang, Y., Western, D., Liu, M., et al. (2024). CSF proteomics identifies early changes in autosomal dominant Alzheimer’s disease. Cell 187, 6309–6326 e6315. 10.1016/j.cell.2024.08.049.

34. Choi, S.R., Golding, G., Zhuang, Z., Zhang, W., Lim, N., Hefti, F., Benedum, T.E., Kilbourn, M.R., Skovronsky, D., and Kung, H.F. (2009). Preclinical properties of 18F-AV-45: a PET agent for Abeta plaques in the brain. J Nucl Med 50, 1887–1894. 10.2967/jnumed.109.065284.

35. Clark, C.M., Schneider, J.A., Bedell, B.J., Beach, T.G., Bilker, W.B., Mintun, M.A., Pontecorvo, M.J., Hefti, F., Carpenter, A.P., Flitter, M.L., et al. (2011). Use of florbetapir-PET for imaging beta-amyloid pathology. JAMA 305, 275–283. 10.1001/jama.2010.2008.

36. Haque, R., Watson, C.M., Liu, J., Carter, E.K., Duong, D.M., Lah, J.J., Wingo, A.P., Roberts, B.R., Johnson, E.C.B., Saykin, A.J., et al. (2023). A protein panel in cerebrospinal fluid for diagnostic and predictive assessment of Alzheimer’s disease. Sci Transl Med 15, eadg4122. 10.1126/scitranslmed.adg4122.

37. Pichet Binette, A., Gaiteri, C., Wennstrom, M., Kumar, A., Hristovska, I., Spotorno, N., Salvado, G., Strandberg, O., Mathys, H., Tsai, L.H., et al. (2024). Proteomic changes in Alzheimer’s disease associated with progressive Abeta plaque and tau tangle pathologies. Nat Neurosci 27, 1880–1891. 10.1038/s41593-024-01737-w.

38. Hristovska, I., Binette, A.P., Kumar, A., Gaiteri, C., Karlsson, L., Strandberg, O., Janelidze, S., van Westen, D., Stomrud, E., Palmqvist, S., et al. (2024). Identification of distinct and shared biomarker panels in different manifestations of cerebral small vessel disease through proteomic profiling. medRxiv. 10.1101/2024.06.10.24308599.

39. Green, G.S., Fujita, M., Yang, H.S., Taga, M., Cain, A., McCabe, C., Comandante-Lou, N., White, C.C., Schmidtner, A.K., Zeng, L., et al. (2024). Cellular communities reveal trajectories of brain ageing and Alzheimer’s disease. Nature 633, 634–645. 10.1038/s41586-024-07871-6.

40. Kelenyi, G. (1967). Thioflavin S fluorescent and Congo red anisotropic stainings in the histologic demonstration of amyloid. Acta Neuropathol 7, 336–348. 10.1007/BF00688089.

41. Urbanc, B., Cruz, L., Le, R., Sanders, J., Ashe, K.H., Duff, K., Stanley, H.E., Irizarry, M.C., and Hyman, B.T. (2002). Neurotoxic effects of thioflavin S-positive amyloid deposits in transgenic mice and Alzheimer’s disease. Proc Natl Acad Sci U S A 99, 13990–13995. 10.1073/pnas.222433299.

42. Levites, Y., Dammer, E.B., Ran, Y., Tsering, W., Duong, D., Abreha, M., Gadhavi, J., Lolo, K., Trejo-Lopez, J., Phillips, J., et al. (2024). Integrative proteomics identifies a conserved Abeta amyloid responsome, novel plaque proteins, and pathology modifiers in Alzheimer’s disease. Cell Rep Med 5, 101669. 10.1016/j.xcrm.2024.101669.

43. Bellenguez, C., Kucukali, F., Jansen, I.E., Kleineidam, L., Moreno-Grau, S., Amin, N., Naj, A.C., Campos-Martin, R., Grenier-Boley, B., Andrade, V., et al. (2022). New insights into the genetic etiology of Alzheimer’s disease and related dementias. Nat Genet 54, 412–436. 10.1038/s41588-022-01024-z.

44. Wingo, A.P., Liu, Y., Mei, Z., Wang, M., Vattathil, S.M., Gerasimov, E.S., Shantaraman, A., Wu, F., Duong, D.M., Fox, E.J., et al. (2025). CSF proteogenomics implicates novel proteins and humoral immunity in Alzheimer’s disease risk. medRxiv, 2025.2009.2021.25336297. 10.1101/2025.09.21.25336297.

45. Persyn, E., Hanscombe, K.B., Howson, J.M.M., Lewis, C.M., Traylor, M., and Markus, H.S. (2020). Genome-wide association study of MRI markers of cerebral small vessel disease in 42,310 participants. Nat Commun 11, 2175. 10.1038/s41467-020-15932-3.

46. Beaufort, N., Scharrer, E., Kremmer, E., Lux, V., Ehrmann, M., Huber, R., Houlden, H., Werring, D., Haffner, C., and Dichgans, M. (2014). Cerebral small vessel disease-related protease HtrA1 processes latent TGF-beta binding protein 1 and facilitates TGF-beta signaling. Proc Natl Acad Sci U S A 111, 16496–16501. 10.1073/pnas.1418087111.

47. Marini, S., Anderson, C.D., and Rosand, J. (2020). Genetics of Cerebral Small Vessel Disease. Stroke 51, 12–20. 10.1161/STROKEAHA.119.024151.

48. Todorovic, V., and Rifkin, D.B. (2012). LTBPs, more than just an escort service. J Cell Biochem 113, 410–418. 10.1002/jcb.23385.

49. Bossi, F., Tripodo, C., Rizzi, L., Bulla, R., Agostinis, C., Guarnotta, C., Munaut, C., Baldassarre, G., Papa, G., Zorzet, S., et al. (2014). C1q as a unique player in angiogenesis with therapeutic implication in wound healing. Proc Natl Acad Sci U S A 111, 4209–4214. 10.1073/pnas.1311968111.

50. Chen, I.H., Wang, H.H., Hsieh, Y.S., Huang, W.C., Yeh, H.I., and Chuang, Y.J. (2013). PRSS23 is essential for the Snail-dependent endothelial-to-mesenchymal transition during valvulogenesis in zebrafish. Cardiovasc Res 97, 443–453. 10.1093/cvr/cvs355.

51. Xue, Q., Wang, X., Deng, X., Huang, Y., and Tian, W. (2020). CEMIP regulates the proliferation and migration of vascular smooth muscle cells in atherosclerosis through the WNT-beta-catenin signaling pathway. Biochem Cell Biol 98, 249–257. 10.1139/bcb-2019-0249.

52. Deroyer, C., Charlier, E., Neuville, S., Malaise, O., Gillet, P., Kurth, W., Chariot, A., Malaise, M., and de Seny, D. (2019). CEMIP (KIAA1199) induces a fibrosis-like process in osteoarthritic chondrocytes. Cell Death Dis 10, 103. 10.1038/s41419-019-1377-8.

53. Fonck, E., Feigl, G.G., Fasel, J., Sage, D., Unser, M., Rufenacht, D.A., and Stergiopulos, N. (2009). Effect of aging on elastin functionality in human cerebral arteries. Stroke 40, 2552–2556. 10.1161/STROKEAHA.108.528091.

54. Duca, L., Blaise, S., Romier, B., Laffargue, M., Gayral, S., El Btaouri, H., Kawecki, C., Guillot, A., Martiny, L., Debelle, L., and Maurice, P. (2016). Matrix ageing and vascular impacts: focus on elastin fragmentation. Cardiovasc Res 110, 298–308. 10.1093/cvr/cvw061.

55. Western, D., Timsina, J., Wang, L., Wang, C., Yang, C., Phillips, B., Wang, Y., Liu, M., Ali, M., Beric, A., et al. (2024). Proteogenomic analysis of human cerebrospinal fluid identifies neurologically relevant regulation and implicates causal proteins for Alzheimer’s disease. Nat Genet 56, 2672–2684. 10.1038/s41588-024-01972-8.

56. Wingo, A.P., Liu, Y., Gerasimov, E.S., Gockley, J., Logsdon, B.A., Duong, D.M., Dammer, E.B., Robins, C., Beach, T.G., Reiman, E.M., et al. (2021). Integrating human brain proteomes with genome-wide association data implicates new proteins in Alzheimer’s disease pathogenesis. Nat Genet 53, 143–146. 10.1038/s41588-020-00773-z.

57. Abraham, H.M., Wolfson, L., Moscufo, N., Guttmann, C.R., Kaplan, R.F., and White, W.B. (2016). Cardiovascular risk factors and small vessel disease of the brain: Blood pressure, white matter lesions, and functional decline in older persons. J Cereb Blood Flow Metab 36, 132–142. 10.1038/jcbfm.2015.121.

58. Zeller, T., Schurmann, C., Schramm, K., Muller, C., Kwon, S., Wild, P.S., Teumer, A., Herrington, D., Schillert, A., Iacoviello, L., et al. (2017). Transcriptome-Wide Analysis Identifies Novel Associations With Blood Pressure. Hypertension 70, 743–750. 10.1161/HYPERTENSIONAHA.117.09458.

59. Schweigert, O., Adler, J., Langst, N., Aissi, D., Duque Escobar, J., Tong, T., Muller, C., Tregouet, D.A., Lukowski, R., and Zeller, T. (2021). CRIP1 expression in monocytes related to hypertension. Clin Sci (Lond) 135, 911–924. 10.1042/CS20201372.

60. Ye, X., Song, Q., Zhang, L., Jing, M., Fu, Y., and Yan, W. (2025). Cysteine-rich intestinal protein family: structural overview, functional diversity, and roles in human disease. Cell Death Discov 11, 114. 10.1038/s41420-025-02395-y.

61. J Duque Escobar, B.B., O Schweigert, R Riedel, P Wenzel, T Zeller (2023). CRIP1: Role in the pathogenesis of hypertension by regulating interactions with cytoskeletal proteins. European Heart Journal 44. doi10.1093/eurheartj/ehad655.3277.

62. Gottesman, R.F., Coresh, J., Catellier, D.J., Sharrett, A.R., Rose, K.M., Coker, L.H., Shibata, D.K., Knopman, D.S., Jack, C.R., and Mosley, T.H., Jr. (2010). Blood pressure and white-matter disease progression in a biethnic cohort: Atherosclerosis Risk in Communities (ARIC) study. Stroke 41, 3–8. 10.1161/STROKEAHA.109.566992.

63. Wartolowska, K.A., and Webb, A.J.S. (2021). Midlife blood pressure is associated with the severity of white matter hyperintensities: analysis of the UK Biobank cohort study. Eur Heart J 42, 750–757. 10.1093/eurheartj/ehaa756.

64. Lai, Y., Jiang, C., Du, X., Sang, C., Guo, X., Bai, R., Tang, R., Dong, J., and Ma, C. (2020). Effect of intensive blood pressure control on the prevention of white matter hyperintensity: Systematic review and meta-analysis of randomized trials. J Clin Hypertens (Greenwich) 22, 1968–1973. 10.1111/jch.14030.

65. Dufouil, C., Chalmers, J., Coskun, O., Besancon, V., Bousser, M.G., Guillon, P., MacMahon, S., Mazoyer, B., Neal, B., Woodward, M., et al. (2005). Effects of blood pressure lowering on cerebral white matter hyperintensities in patients with stroke: the PROGRESS (Perindopril Protection Against Recurrent Stroke Study) Magnetic Resonance Imaging Substudy. Circulation 112, 1644–1650. 10.1161/CIRCULATIONAHA.104.501163.

66. Hampel, H., Elhage, A., Cho, M., Apostolova, L.G., Nicoll, J.A.R., and Atri, A. (2023). Amyloid-related imaging abnormalities (ARIA): radiological, biological and clinical characteristics. Brain 146, 4414–4424. 10.1093/brain/awad188.

67. Spuch, C., Ortolano, S., and Navarro, C. (2012). LRP-1 and LRP-2 receptors function in the membrane neuron. Trafficking mechanisms and proteolytic processing in Alzheimer’s disease. Front Physiol 3, 269. 10.3389/fphys.2012.00269.

68. Bell, R.D., Sagare, A.P., Friedman, A.E., Bedi, G.S., Holtzman, D.M., Deane, R., and Zlokovic, B.V. (2007). Transport pathways for clearance of human Alzheimer’s amyloid beta-peptide and apolipoproteins E and J in the mouse central nervous system. J Cereb Blood Flow Metab 27, 909–918. 10.1038/sj.jcbfm.9600419.

69. Tao, C.C., Cheng, K.M., Ma, Y.L., Hsu, W.L., Chen, Y.C., Fuh, J.L., Lee, W.J., Chao, C.C., and Lee, E.H.Y. (2020). Galectin-3 promotes Abeta oligomerization and Abeta toxicity in a mouse model of Alzheimer’s disease. Cell Death Differ 27, 192–209. 10.1038/s41418-019-0348-z.

70. Seki, T., Kanagawa, M., Kobayashi, K., Kowa, H., Yahata, N., Maruyama, K., Iwata, N., Inoue, H., and Toda, T. (2020). Galectin 3-binding protein suppresses amyloid-beta production by modulating beta-cleavage of amyloid precursor protein. J Biol Chem 295, 3678–3691. 10.1074/jbc.RA119.008703.

71. Garcia-Revilla, J., Boza-Serrano, A., Espinosa-Oliva, A.M., Soto, M.S., Deierborg, T., Ruiz, R., de Pablos, R.M., Burguillos, M.A., and Venero, J.L. (2022). Galectin-3, a rising star in modulating microglia activation under conditions of neurodegeneration. Cell Death Dis 13, 628. 10.1038/s41419-022-05058-3.

72. Kong, A.T., Leprevost, F.V., Avtonomov, D.M., Mellacheruvu, D., and Nesvizhskii, A.I. (2017). MSFragger: ultrafast and comprehensive peptide identification in mass spectrometry-based proteomics. Nat Methods 14, 513–520. 10.1038/nmeth.4256.

73. Yu, F., Haynes, S.E., Teo, G.C., Avtonomov, D.M., Polasky, D.A., and Nesvizhskii, A.I. (2020). Fast Quantitative Analysis of timsTOF PASEF Data with MSFragger and IonQuant. Mol Cell Proteomics 19, 1575–1585. 10.1074/mcp.TIR120.002048.

74. Kall, L., Canterbury, J.D., Weston, J., Noble, W.S., and MacCoss, M.J. (2007). Semi-supervised learning for peptide identification from shotgun proteomics datasets. Nat Methods 4, 923–925. 10.1038/nmeth1113.

75. Yang, K.L., Yu, F., Teo, G.C., Li, K., Demichev, V., Ralser, M., and Nesvizhskii, A.I. (2023). MSBooster: improving peptide identification rates using deep learning-based features. Nat Commun 14, 4539. 10.1038/s41467-023-40129-9.

76. Dammer, E.B., Seyfried, N.T., and Johnson, E.C.B. (2023). Batch Correction and Harmonization of -Omics Datasets with a Tunable Median Polish of Ratio. Front Syst Biol 3. 10.3389/fsysb.2023.1092341.

77. Cai, M., Yue, M., Chen, T., Liu, J., Forno, E., Lu, X., Billiar, T., Celedon, J., McKennan, C., Chen, W., and Wang, J. (2022). Robust and accurate estimation of cellular fraction from tissue omics data via ensemble deconvolution. Bioinformatics 38, 3004–3010. 10.1093/bioinformatics/btac279.

78. Bennett, D.A., Buchman, A.S., Boyle, P.A., Barnes, L.L., Wilson, R.S., and Schneider, J.A. (2018). Religious Orders Study and Rush Memory and Aging Project. J Alzheimers Dis 64, S161–S189. 10.3233/JAD-179939.

79. Barnes, L.L., Shah, R.C., Aggarwal, N.T., Bennett, D.A., and Schneider, J.A. (2012). The Minority Aging Research Study: ongoing efforts to obtain brain donation in African Americans without dementia. Curr Alzheimer Res 9, 734–745. 10.2174/156720512801322627.

80. Marquez, D.X., Glover, C.M., Lamar, M., Leurgans, S.E., Shah, R.C., Barnes, L.L., Aggarwal, N.T., Buchman, A.S., and Bennett, D.A. (2020). Representation of Older Latinxs in Cohort Studies at the Rush Alzheimer’s Disease Center. Neuroepidemiology 54, 404–418. 10.1159/000509626.

81. Love, S., Chalmers, K., Ince, P., Esiri, M., Attems, J., Jellinger, K., Yamada, M., McCarron, M., Minett, T., Matthews, F., et al. (2014). Development, appraisal, validation and implementation of a consensus protocol for the assessment of cerebral amyloid angiopathy in post-mortem brain tissue. Am J Neurodegener Dis 3, 19–32.

82. Kapasi, A., Poirier, J., Hedayat, A., Scherlek, A., Mondal, S., Wu, T., Gibbons, J., Barnes, L.L., Bennett, D.A., Leurgans, S.E., and Schneider, J.A. (2023). High-throughput digital quantification of Alzheimer disease pathology and associated infrastructure in large autopsy studies. J Neuropathol Exp Neurol 82, 976–986. 10.1093/jnen/nlad086.

83. Higginbotham, L., Carter, E.K., Dammer, E.B., Haque, R.U., Johnson, E.C.B., Duong, D.M., Yin, L., De Jager, P.L., Bennett, D.A., Felsky, D., et al. (2023). Unbiased classification of the elderly human brain proteome resolves distinct clinical and pathophysiological subtypes of cognitive impairment. Neurobiol Dis 186, 106286. 10.1016/j.nbd.2023.106286.

84. Reddy, J.S., Heath, L., Linden, A.V., Allen, M., Lopes, K.P., Seifar, F., Wang, E., Ma, Y., Poehlman, W.L., Quicksall, Z.S., et al. (2024). Bridging the gap: Multi-omics profiling of brain tissue in Alzheimer’s disease and older controls in multi-ethnic populations. Alzheimers Dement 20, 7174–7192. 10.1002/alz.14208.

85. Petersen, R.C., Aisen, P.S., Beckett, L.A., Donohue, M.C., Gamst, A.C., Harvey, D.J., Jack, C.R., Jr., Jagust, W.J., Shaw, L.M., Toga, A.W., et al. (2010). Alzheimer’s Disease Neuroimaging Initiative (ADNI): clinical characterization. Neurology 74, 201–209. 10.1212/WNL.0b013e3181cb3e25.

86. Shaw, L.M., Korecka, M., Lee, E.B., Cousins, K.A.Q., Vanderstichele, H., Schindler, S.E., Tosun, D., DeMarco, M.L., Brylska, M., Wan, Y., et al. (2025). ADNI Biomarker Core: A review of progress since 2004 and future challenges. Alzheimers Dement 21, e14264. 10.1002/alz.14264.

87. Chiang, G.C., Cruz Hernandez, J.C., Kantarci, K., Jack, C.R., Jr., Weiner, M.W., and Alzheimer’s Disease Neuroimaging, I. (2015). Cerebral Microbleeds, CSF p-Tau, and Cognitive Decline: Significance of Anatomic Distribution. AJNR Am J Neuroradiol 36, 1635–1641. 10.3174/ajnr.A4351.

88. Fletcher, E., Carmichael, O., and Decarli, C. (2012). MRI non-uniformity correction through interleaved bias estimation and B-spline deformation with a template. Annu Int Conf IEEE Eng Med Biol Soc 2012, 106–109. 10.1109/EMBC.2012.6345882.

89. DeCarli, C., Murphy, D.G., Teichberg, D., Campbell, G., and Sobering, G.S. (1996). Local histogram correction of MRI spatially dependent image pixel intensity nonuniformity. J Magn Reson Imaging 6, 519–528. 10.1002/jmri.1880060316.

90. DeCarli, C., Miller, B.L., Swan, G.E., Reed, T., Wolf, P.A., Garner, J., Jack, L., and Carmelli, D. (1999). Predictors of brain morphology for the men of the NHLBI twin study. Stroke 30, 529–536. 10.1161/01.str.30.3.529.

91. Rajapakse, J.C., Giedd, J.N., DeCarli, C., Snell, J.W., McLaughlin, A., Vauss, Y.C., Krain, A.L., Hamburger, S., and Rapoport, J.L. (1996). A technique for single-channel MR brain tissue segmentation: application to a pediatric sample. Magn Reson Imaging 14, 1053–1065. 10.1016/s0730-725x(96)00113-0.

92. Fletcher, E., Singh, B., Harvey, D., Carmichael, O., and DeCarli, C. (2012). Adaptive image segmentation for robust measurement of longitudinal brain tissue change. Annu Int Conf IEEE Eng Med Biol Soc 2012, 5319–5322. 10.1109/EMBC.2012.6347195.

93. Kueper, J.K., Speechley, M., and Montero-Odasso, M. (2018). The Alzheimer’s Disease Assessment Scale-Cognitive Subscale (ADAS-Cog): Modifications and Responsiveness in Pre-Dementia Populations. A Narrative Review. J Alzheimers Dis 63, 423–444. 10.3233/JAD-170991.

94. Chawla, N.V.B., K.W.; Hall, L.O.; Kegelmeyer, W.P. (2002). SMOTE: Synthetic Minority Over-sampling Technique. Journal of Artificial Intelligence Research 16. 10.1613/jair.953.

95. Guyon, I., Weston, J., Barnhill, S., Vapnik V. (2002). Gene Selection for Cancer Classification using Support Vector Machines. Machine Learning 46, 389–422. 10.1023/A:1012487302797.

96. Youden, W.J. (1950). Index for rating diagnostic tests. Cancer 3, 32–35. 10.1002/1097-0142(1950)3:1<32::aid-cncr2820030106>3.0.co;2-3.

97. Chang, C.C., Chow, C.C., Tellier, L.C., Vattikuti, S., Purcell, S.M., and Lee, J.J. (2015). Second-generation PLINK: rising to the challenge of larger and richer datasets. Gigascience 4, 7. 10.1186/s13742-015-0047-8.

98. Gusev, A., Ko, A., Shi, H., Bhatia, G., Chung, W., Penninx, B.W., Jansen, R., de Geus, E.J., Boomsma, D.I., Wright, F.A., et al. (2016). Integrative approaches for large-scale transcriptome-wide association studies. Nat Genet 48, 245–252. 10.1038/ng.3506.

99. Zhu, Z., Zhang, F., Hu, H., Bakshi, A., Robinson, M.R., Powell, J.E., Montgomery, G.W., Goddard, M.E., Wray, N.R., Visscher, P.M., and Yang, J. (2016). Integration of summary data from GWAS and eQTL studies predicts complex trait gene targets. Nat Genet 48, 481–487. 10.1038/ng.3538.

